# Behavioral interventions to improve sleep outcomes in people with multiple sclerosis: A systematic review

**DOI:** 10.1101/2022.10.29.22281670

**Authors:** David Turkowitch, Sarah J. Donkers, Silvana L. Costa, Prasanna Vaduvathiriyan, Joy Williams, Catherine Siengsukon

**Affiliations:** Department of Physical Therapy, Rehabilitation Science, and Athletic Training, University of Kansas Medical Center, Kansas City, KS, USA; School of Rehabilitation Science, University of Saskatchewan, Saskatoon, Canada; Center for Neuropsychology and Neuroscience Research, Kessler Foundation, East Hanover, NJ; Department of Physical Medicine and Rehabilitation, Rutgers –New Jersey Medical School, Newark, NJ, USA; A.R. Dykes Library, University of Kansas Medical Center, Kansas City, KS, USA; Rehabilitation Services, Kaiser Permanente Sacramento, Sacramento, CA, USA

**Keywords:** Behavioral Intervention, Multiple Sclerosis, Sleep, Exercise, Physical Activity, CBT-I, CBT, Mindfulness, Education, Complementary Medicine

## Abstract

**Objective:** To determine effective behavioral interventions to improve sleep in people with MS.

**Methods:** Systematic review following PRISMA guidelines.

**Data Sources:** Literature searches were performed in December 2021 in Ovid MEDLINE, CINAHL, and Web of Science along with hand searching for grey literature and cited references. Out of the 837 search results, 830 unique references were reviewed after duplicates were removed.

**Study Selection:** Four reviewers independently reviewed titles and abstracts (two reviewers for each article), and a fifth reviewer resolved discrepancies. The full-text articles (n = 81) were reviewed independently by four reviewers (two for each article) for eligibility, and consensus for inclusion was achieved by a fifth reviewer as needed. Thirty-seven articles were determined eligible for inclusion.

**Data Extraction:** Four reviewers extracted relevant data from each study (two reviewers for each article) using a standard data-extraction table. Consensus was achieved for completeness and accuracy of the data extraction table by a fifth reviewer. Four reviewers (two reviewers for each article) conducted a quality appraisal of each article to assess the risk for bias and quality of the articles and consensus was achieved by a fifth reviewer as needed.

**Data Synthesis:** Descriptions were used to describe types of interventions, sleep outcomes, results, and key components across interventions.

**Conclusions:** The variability in the intervention types, intervention dose, outcomes used, training/expertise of interventionist, specific sample included, and quality of the study made it difficult to compare and synthesize results. Overall, the CBT-I, CBT/psychotherapy, and education/self-management support interventions reported positive improvements in sleep outcomes. The quality appraisal scores ranged from low to high quality indicating potential for bias. Further research is necessary to demonstrate efficacy of most of the interventions.

---

Nearly one million people in the US are living with multiple sclerosis (MS), and in this population, disturbed sleep is a very common challenge.^1-3^ Sleep problems are significantly more common in people with MS than in the general population, estimated at nearly 70% of individuals with MS.^4,5^ However, the incidence is likely higher as sleep disorders in individuals with MS are severely underdiagnosed.^6,7^

Common contributors of sleep disturbances in people with MS include nocturia, muscle spasticity, depression, anxiety, and pain.^8-10^ Furthermore, neuropathology due to MS may have direct physiological impact on the circadian rhythm,^11,12^ and side effects from medications can also have a significant effect on sleep, further complicating the treatment of sleep disturbances.^9^ Sleep disturbances in people with MS have been associated with poorer cognitive performance, lower quality of life, higher disability, and increased prevalence of pain, fatigue, depression, anxiety, and sexual and bladder dysfunction.^13-18^ Because of the overlapping symptoms of MS and disturbed sleep, poor sleep in people with MS is a complicated issue to treat.

Various pharmacological and lifestyle/behavioral interventions have been studied to address sleep disturbances in people with MS. Although pharmacological interventions are appropriate in certain cases, medication can have negative side effects (such as sedation, dizziness, cognitive impairment, and motor incoordination) and concerns for long-term use including exacerbation of sleep disturbances and insomnia.^19-21^ Conversely, behavioral interventions for the treatment of MS are safe for short and long-term use, have shown promising impact on sleep issues, may be more accessible, and may potentially modify the disease process.^22^ Behavioral interventions that have shown effective, durable, and safe results include cognitive behavioral therapy for insomnia (CBT-I), physical activity, mindfulness training, education, and psychotherapy. However, the effective components of sleep interventions in people with MS have yet to be summarized.

Accordingly, this systematic review focuses on the behavioral interventions being implemented to improve sleep in people with MS. The body of research examining behavioral interventions to improve sleep in people with MS is large and rapidly expanding; thus, the literature in the field has been assembled for close examination. The purpose of this systematic review is to determine effective behavioral interventions to improve sleep in people with MS. It is anticipated that this systematic review will act as a guide for clinical practice and future research in this burgeoning field by identifying efficacious behavioral interventions to enhance sleep as well as gaps in knowledge.

## Methods

The Preferred Reporting Items for Systematic Reviews and Meta-Analyses (PRISMA) guidelines were used for identifying the standards of this review protocol. This systematic review was prospective registered at the International Prospective Register of Systematic Reviewers (PROSPERO; ID CRD42022302318). Ovid Medline, Embase.com, and Web of Science were used for identifying relevant studies. Both keywords and database provided controlled vocabularies are used in Embase and Ovid Medline databases along with various search techniques and Boolean operators. The Web of Science database was searched using keywords only. These terms included: multiple sclerosis, sleep, sleep wake disorders, circadian rhythm, exercise, exercise therapy, etc. (refer to supplementary file 1 for detailed search strategies). To include non-pharmaceutical intervention outcome of sleep quality, a wide variety of terms such as behavioral therapy, complementary therapy, mind body therapy, muscle relaxation, qigong, mindfulness, meditation, etc. were included in the search. Publication of language filters were not applied. Additional resources such as ClinialTrials.gov and Google scholar used for identifying relevant grey literature on the topic were also searched. Searches were performed between December 1st and 28^th^, 2021.

Studies were included in the systematic review if they: (1) involved the study of humans diagnosed with Multiple Sclerosis; (2) consisted of an intervention study design (i.e. pre-post test, randomized and non-randomized trial study design); (3) included lifestyle or behavioral interventions; and (4) included sleep as an outcome. Studies were excluded from the systematic review if they: (1) included the study of animals; (2) involved any other neurologic condition; (3) were written in a non-English language; (4) involved irrelevance to objectives; (5) included non-intervention study design (reviews, opinions, editorials, protocols, cross-sectional, observational); and (6) contained a duplicate data set (same data reported in more than one study).

Each title/abstract was independently reviewed by two of the four reviewers (S.L.C., S.J.D., C.S., and D.T) and determined as eligible or ineligible based on the above inclusion/exclusion criteria). Discrepancies were resolved by a fifth reviewer (J.W.) in order to reach consensus. The same four reviewers then reviewed the full-text manuscripts for eligibility, and consensus was reached on discrepancies by the fifth reviewer.

Data was then extracted from the full-text articles by both a primary reviewer and a secondary reviewer (S.L.C., S.J.D., C.S., and D.T) using a standardized data extraction spreadsheet. Information extracted included the first author, publication year, country the study was conducted, study design, study objective, number of participants enrolled, number of dropouts, participant information including age, MS-type, severity, disease duration, specific impairment(s) for eligibility, intervention information including type, dose, interventionist, setting, study assessment timepoints, control group (if included), the primary and sleep outcome(s), and sleep-specific results. Consensus was reached on discrepancies by the fifth reviewer.

To assess the risk for potential bias and quality of the included articles, critical appraisals were independently conducted by four reviewers (two for each article; same reviewers who conducted data extraction), and discrepancies were resolved by the fifth reviewer. Five different quality appraisal tools were used according to article types: National Heart, Lung, and Blood Institute quality assessment for before-after (pre-post) studies with no control group,^23^ and Joanna Briggs Institute’s (JBI) critical appraisal checklists for randomized controlled trials (RCT), case reports, quasi-experimental studies, and qualitative research.^24^ Overall quality scores were then calculated for each study as follows: “yes” = 2, “unclear” and “cannot determine” = 1, and “not reported” or “no” = 0, and a percentage score was calculated.^25^ A score of 100% would indicate the highest possible quality score and demonstrate the quality in study design and implementation to reduce potential bias. Because the twelfth question was rated as “not applicable” for all the pre-post study design articles, the denominator was adjusted to eleven X 2 rather than twelve X 2 to calculate overall percent quality score. Also, two questions in the case series quality appraisal were deemed “not applicable” for the case series, so the denominator was adjusted to eight X 2 rather than ten X 2. In order to be consistent with other checklists, a “not reported” option to the JBI RCT checklist was added. Only one reviewer conducted quality appraisal for the qualitative research studies due to her being the sole reviewer with expertise in qualitative design. Also, since there is not a specific checklist for pilot RCTs, the JBI RCT appraisal checklist was used for studies that self-identified as a pilot RCT.

## Results

The search strategy generated 1,269 total citations. There were 439 duplicates identified and removed, leaving 839 unique results. Subsequent title/abstract review excluded 749 studies, leaving 81 studies for full-text review. Following full text review, 44 studies were excluded, leaving 37 studies for data extraction and quality appraisal (Figure 1).

**Figure 1.**
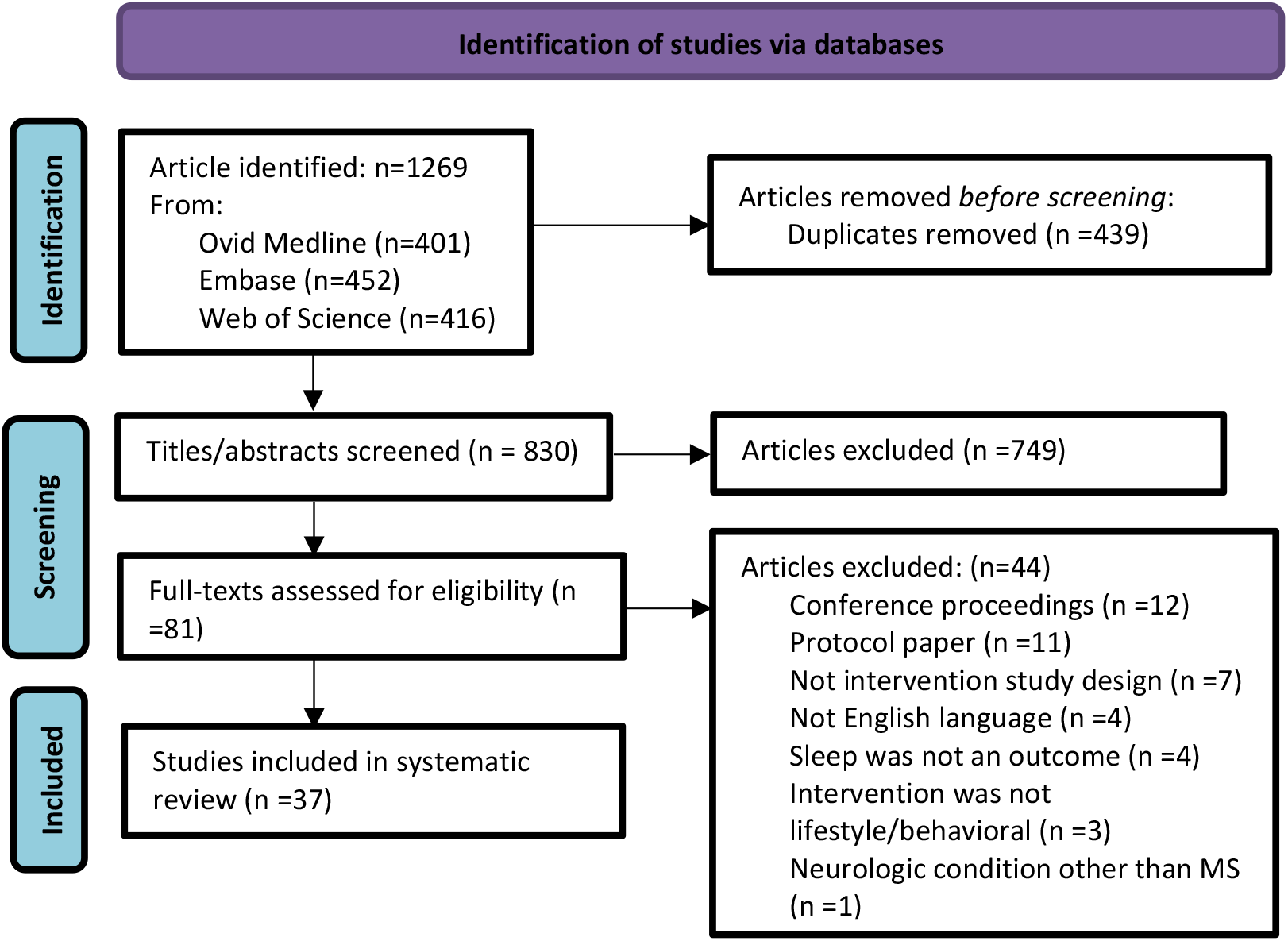
PRISMA flow diagram.

The studies that met the inclusion/exclusion criteria for this systematic review are summarized in Table 1 (organized by intervention type and by study design). Fifty one percent of the studies were conducted in North America (94% of which were in the United States), 29% in Europe, 23% in Asia, and 3% in Australia. The most prevalent study design was RCT (n=19^26-35^; 54%; of which n=9^36-44^ self-identified as being a pilot RCT) followed by single group pre-post design (n=11^45-55^; 31%). Case reports (n=2^56,57^; 6%), qualitative studies (n=2^58,59^; 6%), quasi experimental studies (n=2^60,61^; 6%), and case series (n=1^62^; 3%), were also included. Four of the studies included are secondary analyses,^27,58,59,63^ two of which^59,63^ analyze data from a primary study also included in this systematic review.^42,60^

**Table 1.**
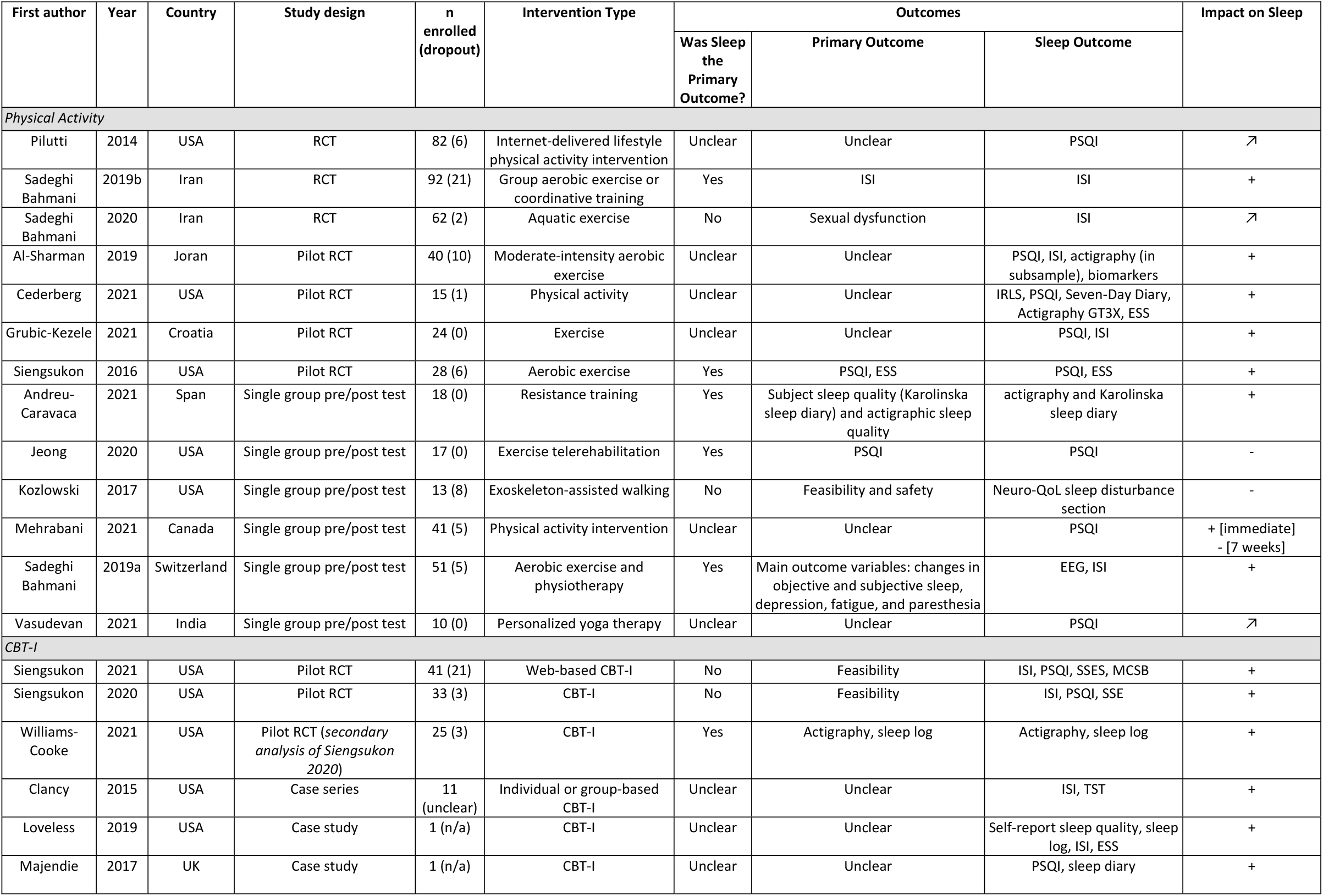
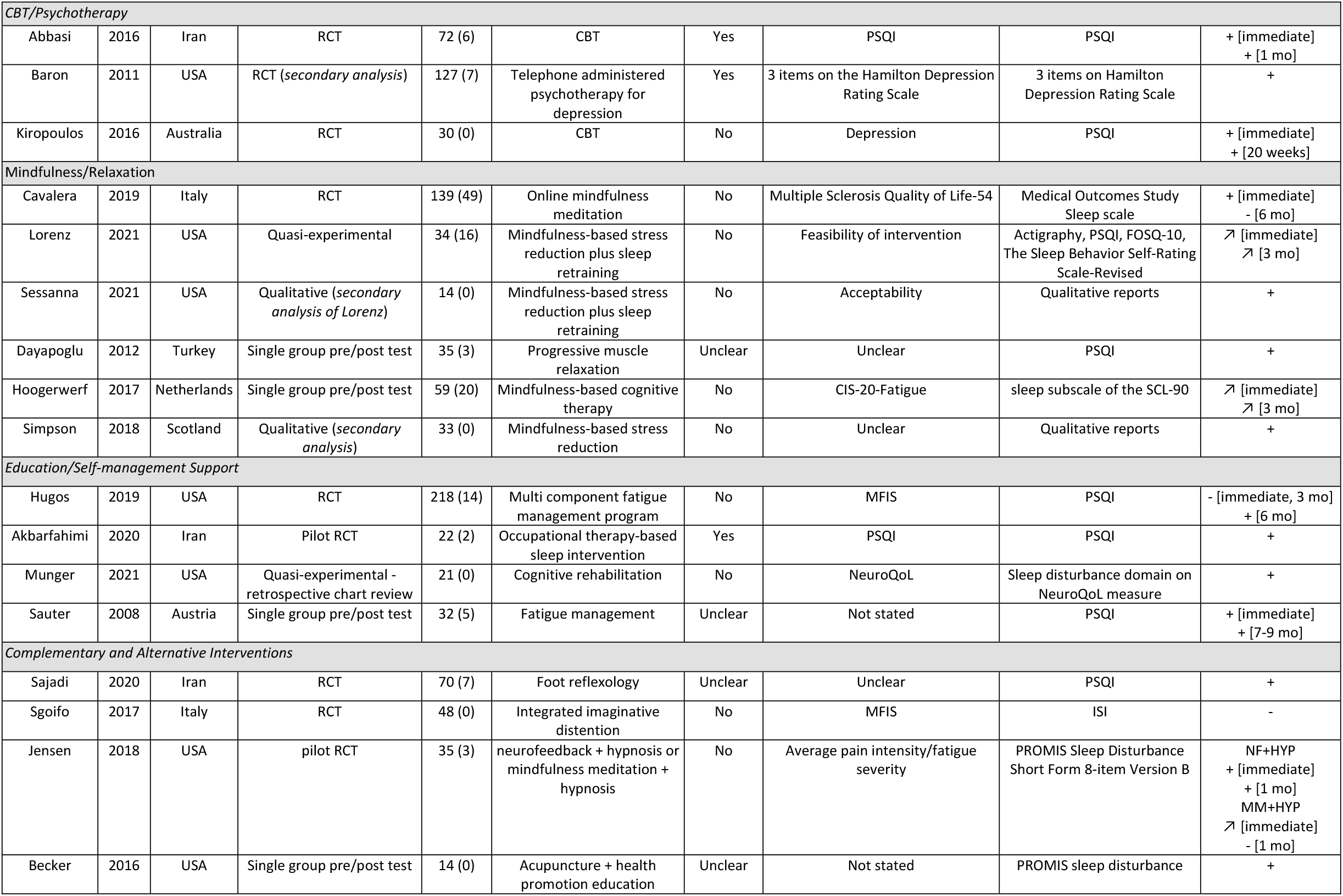
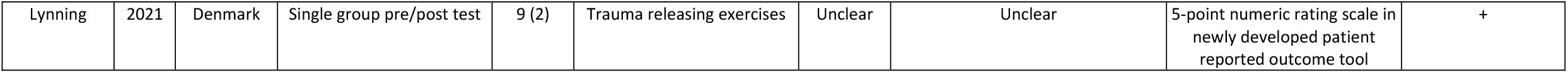
Summary of articles. + = improvement in sleep; - = worsening, no improvement in sleep, or mixed results; ↗ = Improvement not statistically significant; FOSQ-10 = Functional Outcomes of Sleep Questionnaire; MCSB = Motivation to Change Sleep Behavior; SSES = Sleep Self-efficacy Scale; TST = total sleep time

Sample sizes ranged from one^56,57^ to 218 individuals^29^, and 1,565 individuals were included in total. Participants in all of the studies included were adults ranging in age from 34.3 years old^26^ to 60.1 years old.^50^ Relapse-remitting MS constituted the majority of MS types for every study except for three studies.^39,51,57^ Sleep was stated as the primary outcome in nine^26,27,32,36,41,43,45,46,50^ of the 37 studies. The Pittsburg Sleep Quality Index (PSQI) was the most commonly used sleep outcome utilized in 19^26,29-31,35-39,41,42,44,48,50,53-55,57,60^ of the studies followed by the Insomnia Severity Index (ISI) (n=10^32-34,37,39,42,44,45,56,62^), actigraphy (n=5^37,38,43,46,60^), and sleep log/diary data (n=5^38,43,46,56,57^); six of the studies incorporated an objective measurement (actigraphy (n=5^38,43,46,56,57^); EEG (n=1^45^)). Interventions included physical activity (n=13^31-33,37-39,41,45,46,50,51,53,55^), CBT-I (n=6^42-44,56,57,62^), CBT/psychotherapy (n=4^26,27,30,^), mindfulness/relaxation (n=6^28,48,49,58-60^), education and self-management support (n=3^29,36,54,61^), and complementary and alternative interventions including acupuncture^47^, neurofeedback and hypnosis^64^, trauma releasing exercises^52^, foot reflexology^35^, integrated imaginative distention^34^ (all n=1). Details regarding sample demographics, intervention provided, and the major sleep findings are summarized by intervention category in Table 2. Results of studies will next be summarized and described by intervention category.

**Table 2.**
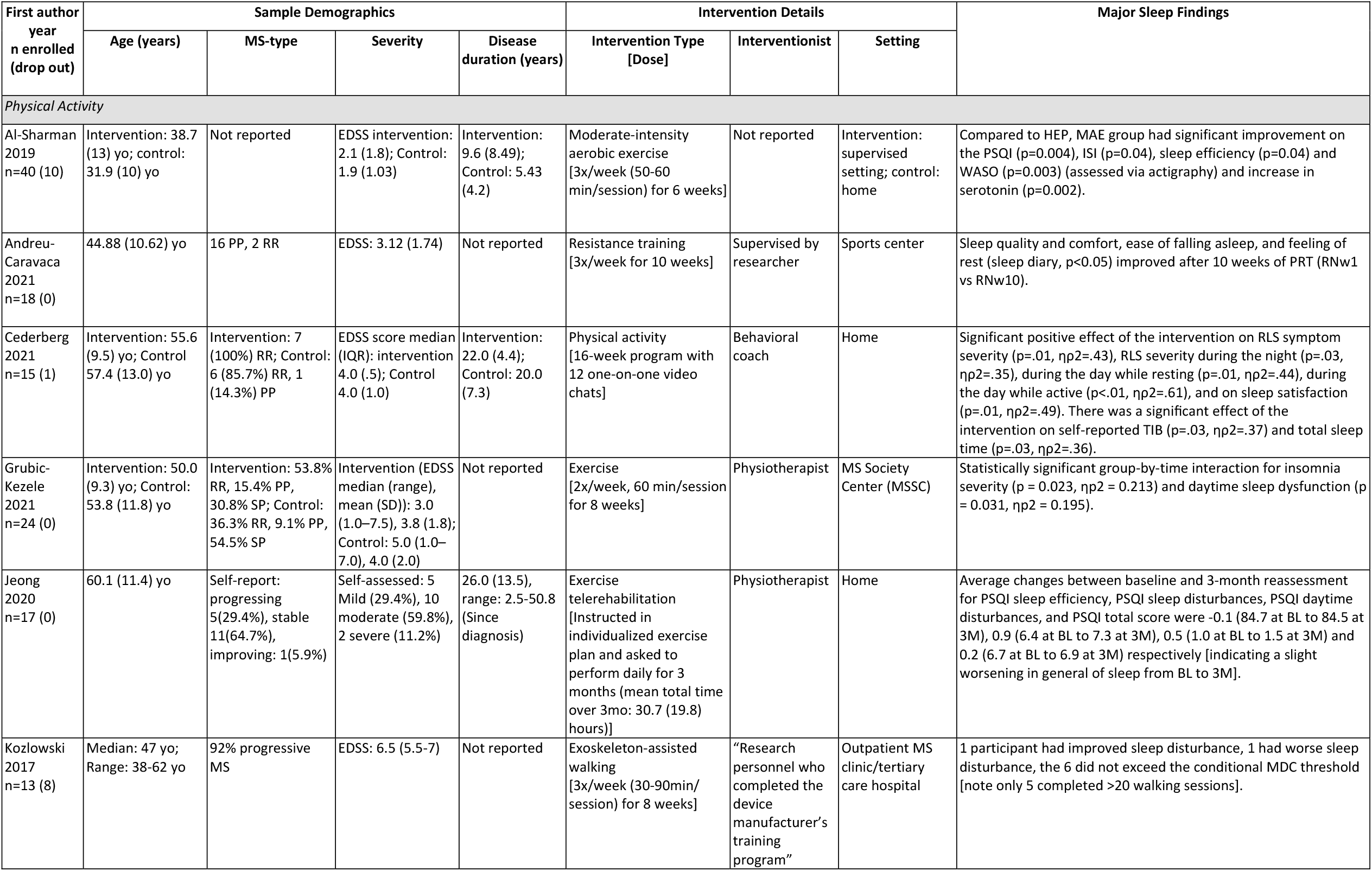
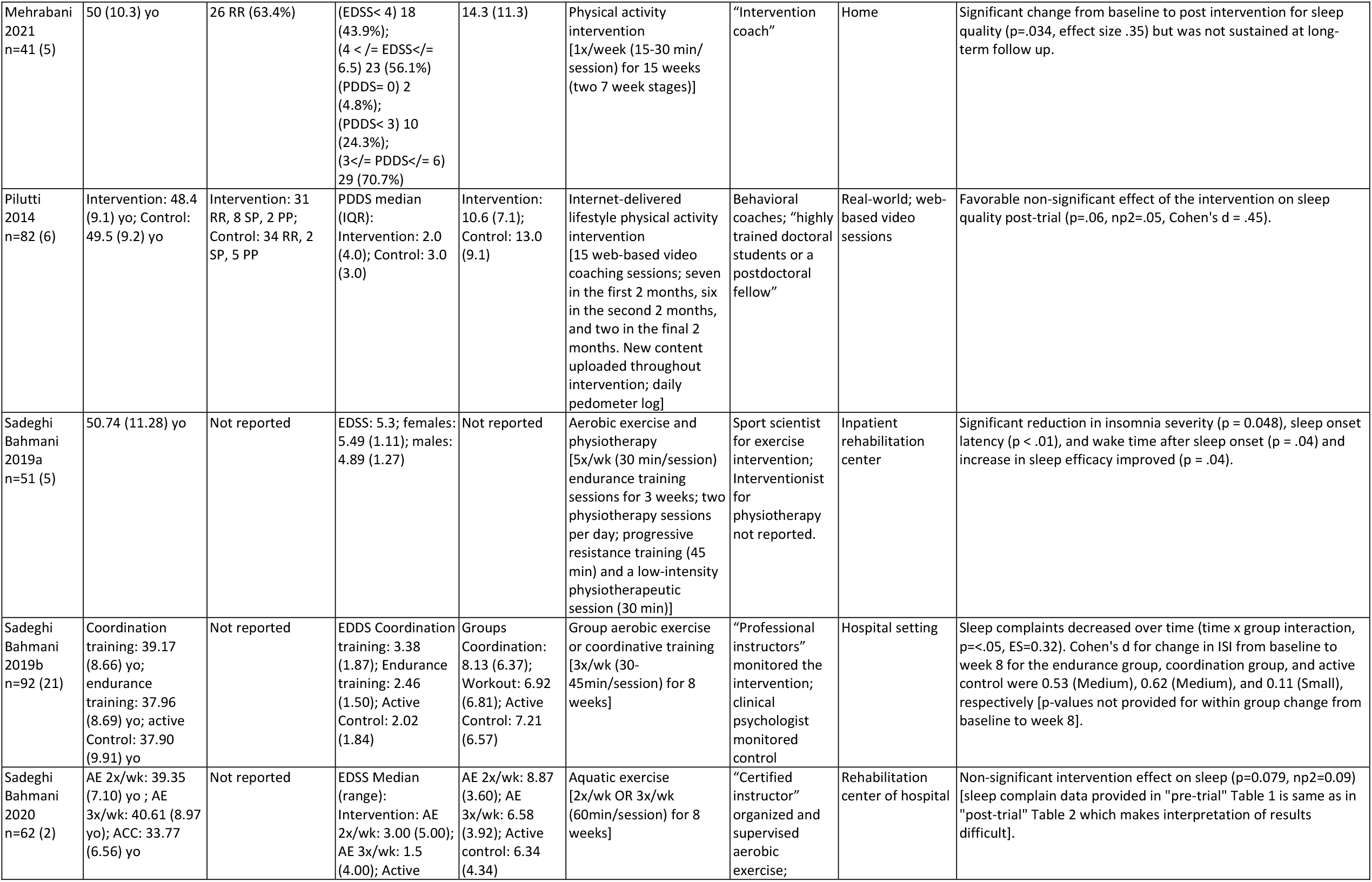
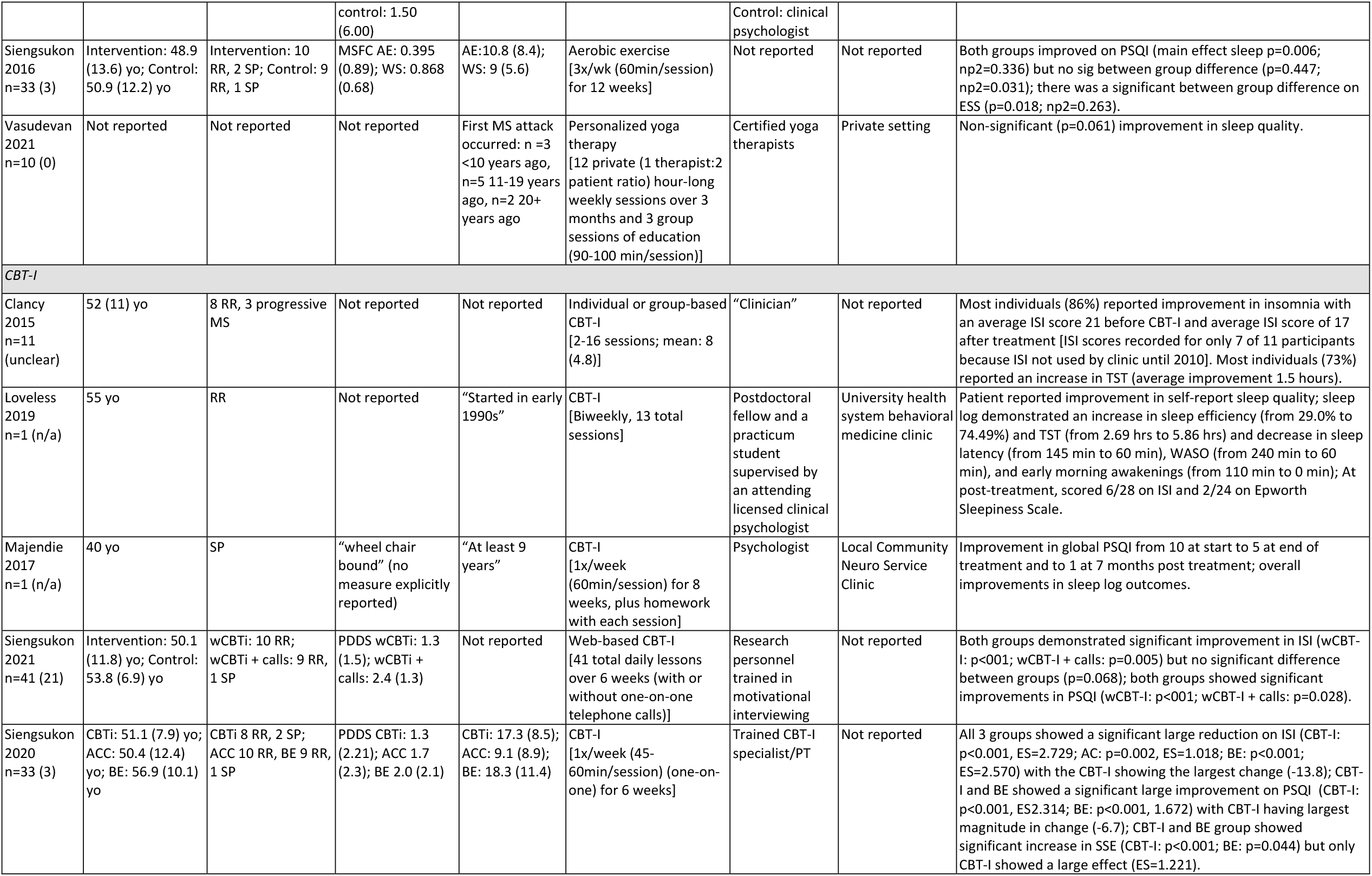
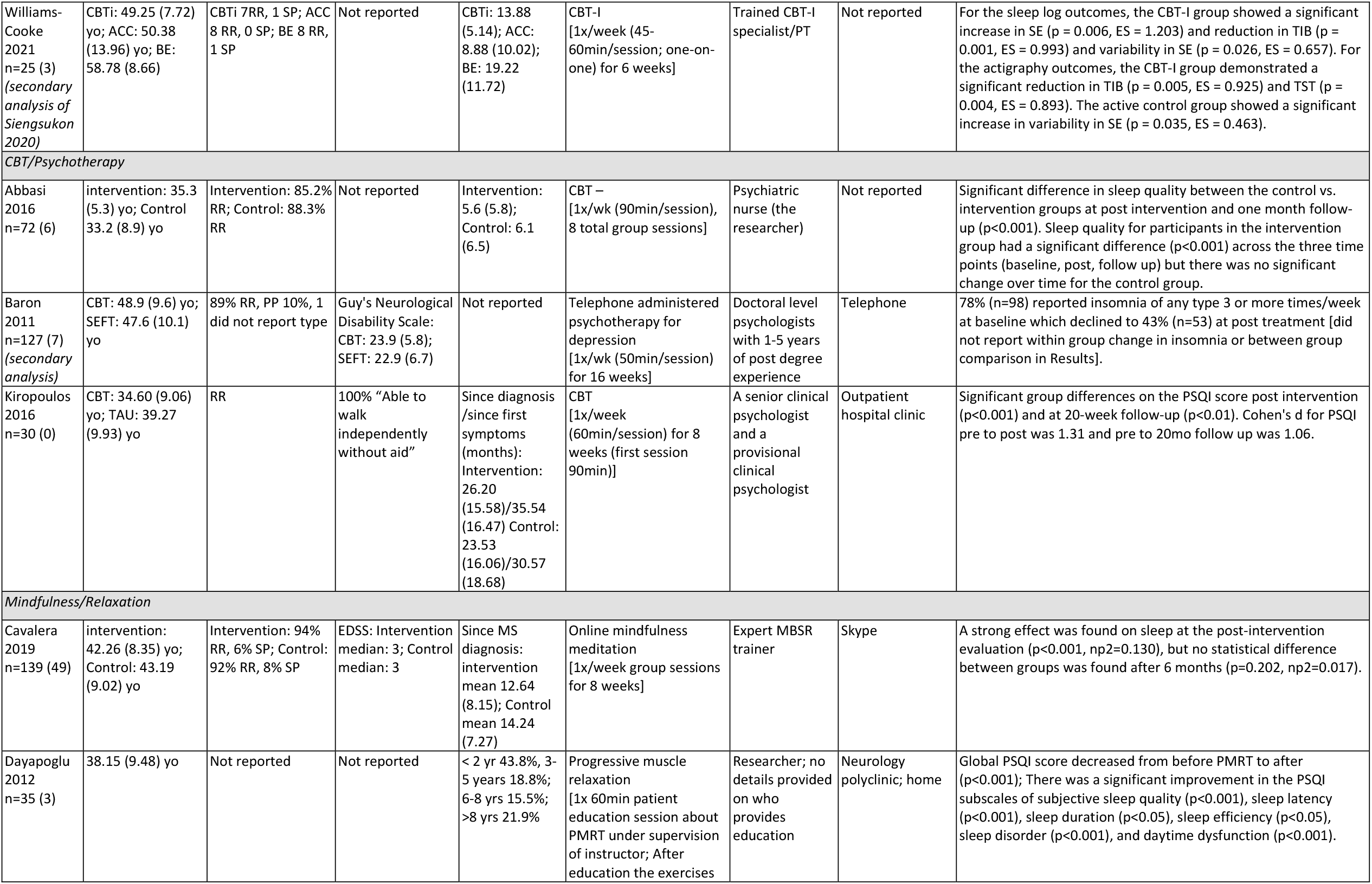
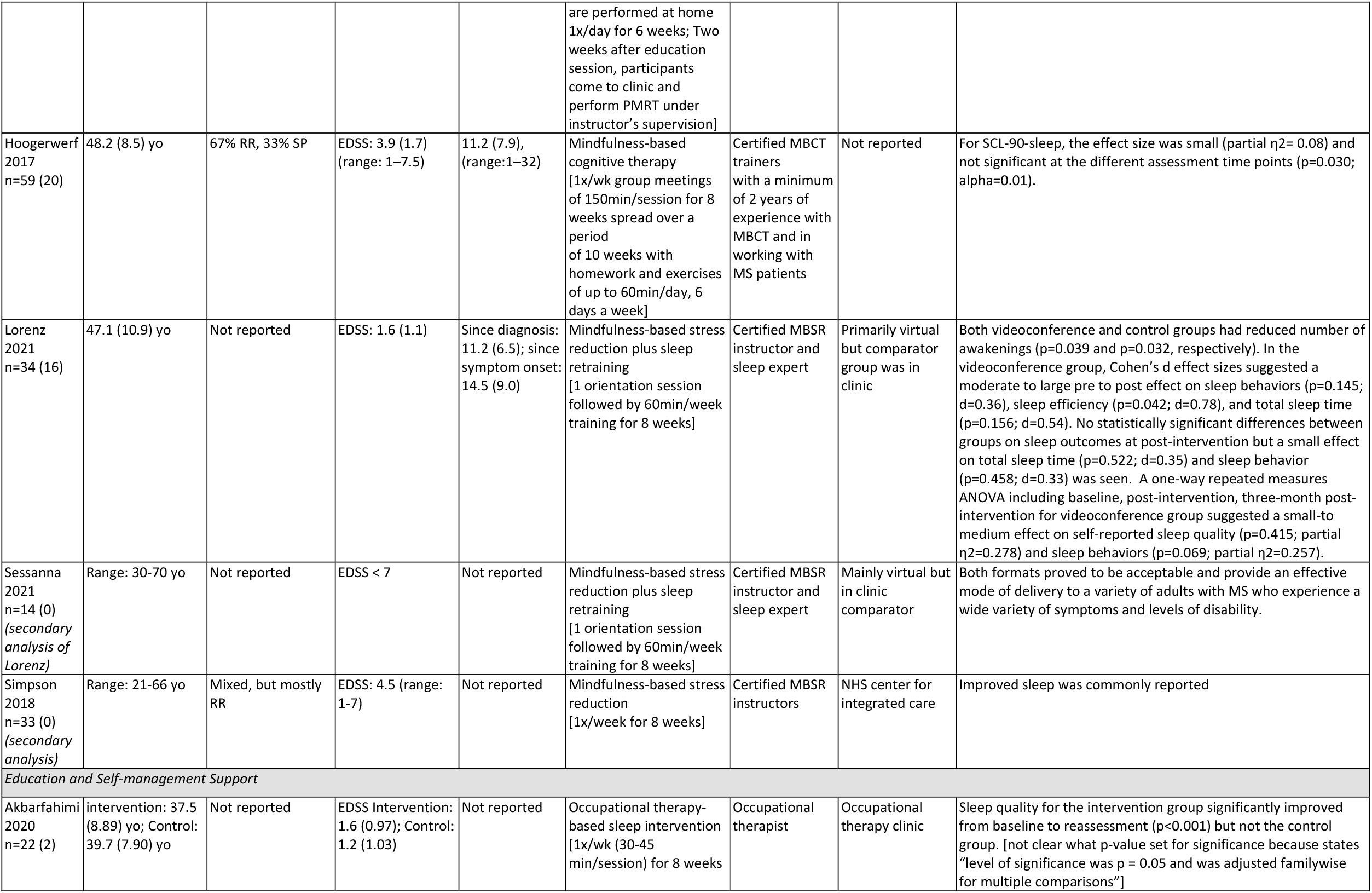
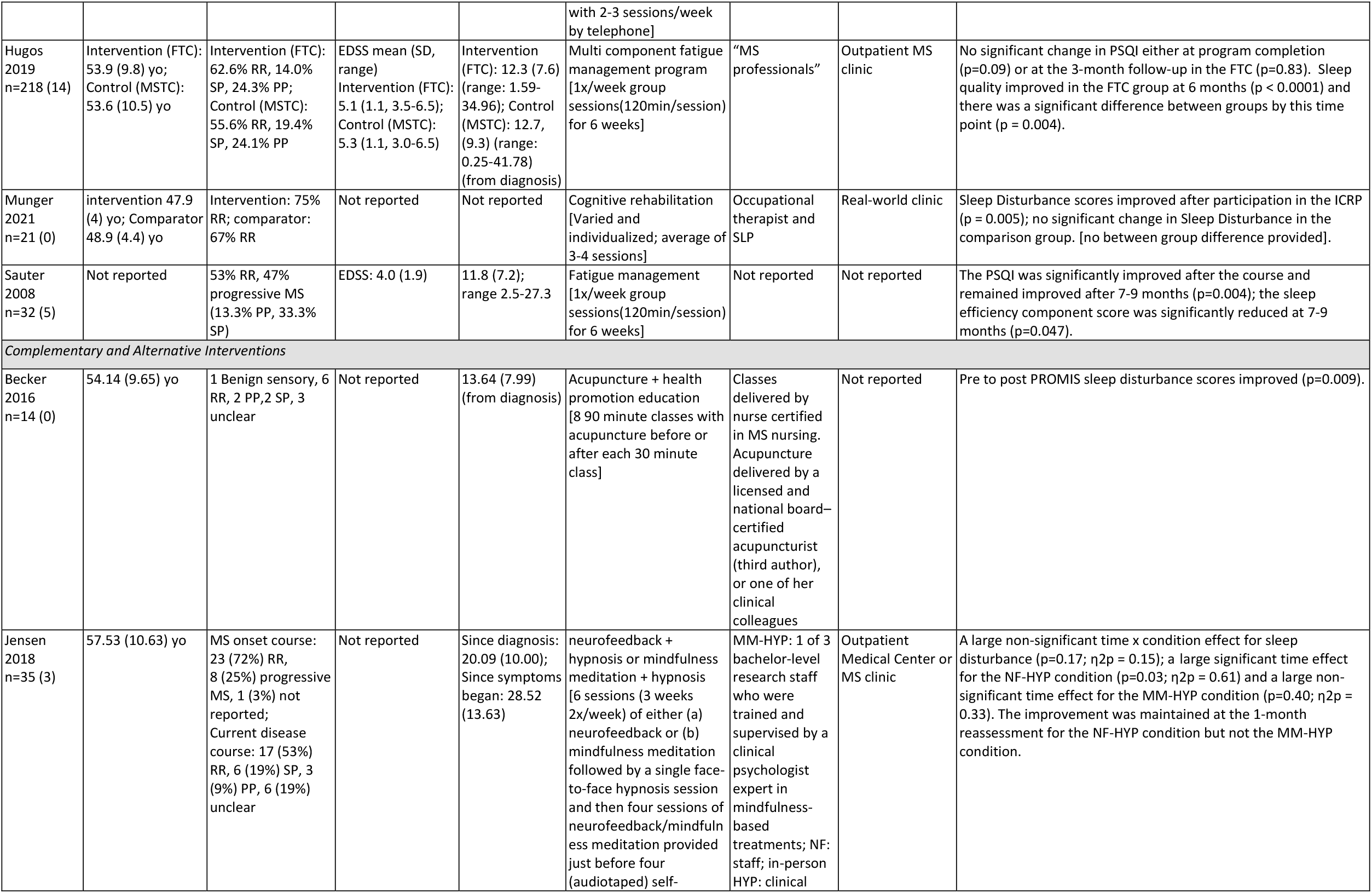
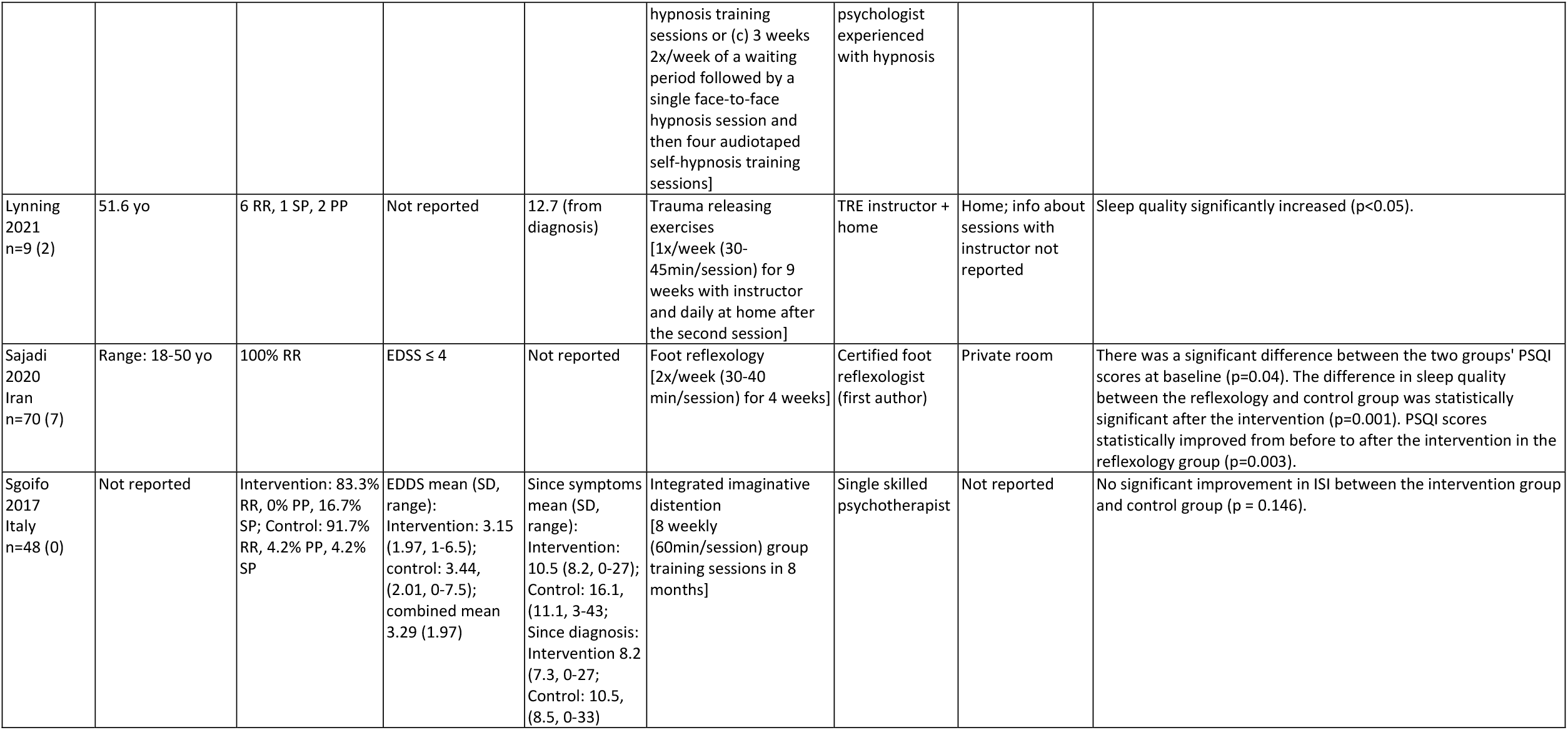
Details of study sample, intervention, and major sleep finding. 3M = 3 month reassessment; ACC = Active Control Condition; AE = Aquatic/Aerobic Exercise; BE = Basic Education; Bl = baseline; EDSS = Expanded Disease Status Scale; FTC = Fatigue: Take Control; HEP = Home Exercise Program; HYP = Hypnosis; IAYT = International Association of Yoga Therapists; ICRP= Integrated Cognitive Rehabilitation Program; MAE = Moderate-Intensity Aerobic Exercise Program; MBCT = Mindfulness Based Cognitive Therapy; MBSR = Mindfulness Based Stress Reduction; MDC = minimal detectable change; MCID = Minimal Clinically Important Difference; MFIS = Modified Fatigue Impact Scale; MM = Mindfulness Meditation; MSFC = Multiple Sclerosis Functional Composite; MSTC = Multiple Sclerosis: Take Control; NHS = National Health Service; PDDS = Patient Determined Disease Steps; PHQ-9 = Patient Health Questionnaire; PMRT = Progressive Muscle Relaxation Technique; PP = Primary Progressive [Multiple Sclerosis]; PT = Physical Therapist; RLS = Restless Legs Syndrome; RN = Rest Night; RR = Relapse-Remitting [Multiple Sclerosis]; SE = Sleep Efficiency; SEFT = Supportive Emotion-Focused Therapy; SLP = Speech-Language Pathologist; SP = Secondary Progressive [Multiple Sclerosis]; SSE = Sleep Self Efficacy; TAU = Treatment as Usual; TIB = Time in Bed; TN = Training Night; TRE = Trauma Releasing Exercises; TST = Total Sleep Time; WS = Walking and Stretching program.

### Physical activity

Thirteen of the 37 included studies incorporated physical activity as a behavioral intervention.^31-33,37-39,41,45,46,50,51,53,55^ A variety of physical activity interventions were used including resistance training,^46^ aerobic exercise,^32,37,41,45^ general individualized exercise programs,^39,50^ physical activity lifestyle interventions,^31,38,53^ yoga,^55^ exoskeleton assisted walking,^51^ and aquatic exercise.^33^ Five of the physical activity studies stated sleep was a primary outcome.^32,41,45,46,50^ Of the five studies with sleep as the primary outcome, three were single group pre-post studies^45,46,50^, and two were RCTs^65^ (one being a self-identified pilot RCT^41^).

All four pilot physical activity RCTs had statistically significant improvement in sleep outcomes (effect size ranging from small to large).^37-39,41^ Siengsukon et al.^41^ was the only pilot physical activity RCT in which sleep was explicitly included the primary outcome. Siengsukon et al.^41^ reported that both the 12-week moderate-intensity aerobic exercise group and the low impact walking and stretching group experienced large significant improvements on the PSQI, but there was no significant between group difference; however, a significant between group difference was found on the Epworth Sleepiness Scale (ESS; large effect).

Cederberg & Motl^38^ reported on a sample of individuals with MS and restless legs syndrome. After 16 weeks of a physical activity intervention, they observed significant positive effects on the International Restless Legs Syndrome Study Group Scale (IRLS), restless legs syndrome (RLS) severity during the night and during the day while either resting and while active, sleep satisfaction, self-reported time in bed, and total sleep time following a physical activity intervention compared to the waitlist control. In addition, other sleep outcomes demonstrated moderate to large effect sizes although were not statistically significant.

Al-Sharman et al.^37^ reported that the six-week moderate-intensity aerobic exercise group had significant moderate-to-large improvements on the global PSQI, ISI, sleep efficiency, and wake time after sleep onset assessed via actigraphy compared to a home exercise group. Furthermore, change in serotonin was significantly correlated with change in PSQI within the aerobic exercise group.

Grubic-Kezele et al.^39^ reported a significant large improvement on the ISI following an eight-week individualized exercise program compared to socializing at the MS Society Center. The only PSQI measure that showed a significant large improvement was daytime sleep dysfunction. In addition, other sleep outcomes demonstrated small to large effect sizes although were not statistically significant.

Two of the RCTs reported medium effect sizes for ISI change following eight-week endurance training and coordinative training^65^ and eight-week, 2x/week aquatic exercise group.^33^ However, between group statistical comparisons were not provided in either study. In the third RCT study,^31^ there was a moderate non-significant improvement in PSQI in the intervention group that participated in a 24-week internet-delivered behavioral intervention to increase lifestyle physical activity, and there was no significant between group differences reported.

Three of the six single group pre-post study design physical activity studies stated sleep was a primary outcome. Jeong et al.^50^ reported a slight worsening of PSQI scores from baseline to 3 months of a daily individualized at-home exercise intervention. Sadeghi Bahmani et al.’s study^45^ on three weeks of endurance and physiotherapy sessions reported significantly improved ISI scores, sleep onset latency, sleep efficiency, and wake after sleep onset (WASO). Andreu-Caravaca et al.^46^ demonstrated improvements in sleep efficiency (via actigraphy) and sleep quality, comfort, feeling of rest, and ease of falling asleep (via Karolinska Sleep Diary) following resistance training sessions compared to non-training sessions.

In the other three single group pre-post study design physical activity studies, sleep was not the primary outcome or it was unclear if sleep was a primary outcome. Vasudevan et al.^55^ reported non-significant improvement on the PSQI using a 12-week personalized yoga program. Mehrebani et al.^53^ reported significant improvement in sleep quality immediately after a 15-week physical activity intervention; however, the improvement was not sustained seven weeks post-intervention. Kozlowski et al.’s^51^ eight-week study on exoskeleton assisted walking reported mix results using the Quality of Life in Neurological Conditions (NeuroQoL) sleep disturbance scale.

### CBT-I

Six^42-44,56,57,62^ of the 37 total studies incorporated in this systematic review incorporated a CBT-I intervention, and all reported improved sleep outcomes. Three of these studies were pilot RCTs,^42,44^ one of which was a secondary analysis.^43^ Two of the CBT-I studies were case reports,^56,57^ and one was a case series.^62^ Sleep was stated as the primary outcome in only the secondary analysis.^43^

In Siengsukon et al.’s two pilot RCT studies, both the six-week web-based CBT-I^44^ and six week in-person CBT-I^42^ interventions produced significant large improvements in PSQI, ISI, and SSE scores. Secondary analysis by Williams-Cooke et al.^43^ of the in-person CBT-I study revealed sleep log data with significant moderate-to-large improvements in sleep efficacy and reduction in time in bed and variability in sleep efficacy. Actigraphy data from the CBT-I group demonstrated a significant large reduction in time in bed and total sleep time from baseline to post-intervention and the active control group showed a significant increase in variability in sleep efficiency.

Both individuals involved in the case reports demonstrated improved sleep outcomes. The case report by Loveless et al.^56^ included a mid-50 year old male with relapse remitting MS. After 13 total CBT-I sessions, the patient’s sleep log demonstrated an increase in sleep efficiency (from 29.0% to 74.49%) and total sleep time (from 2.69 to 5.86 hours) and decreases in sleep latency (from 145 to 60 minutes), wake after sleep onset (from 240 to 60 minutes), and early morning awakenings (from 110 min to 0 min). The case report by Majendie et al.^57^ included a 40-year-old male with secondary progressive MS and sleep disturbance. His global PSQI score improved from ten to five following eight sessions of CBT-I and was reported to be a one 7 months post-treatment. Sleep onset latency, sleep duration, and sleep efficiency also improved, and the participant implemented a medication withdrawal plan during his intervention time frame. In the case series by Clancy et al.^62^ most (86%) patients reported improvements in both ISI scores and total sleep time after an average of eight CBT-I sessions.

### CBT/Psychotherapy

Three studies were included in the systematic review that incorporated CBT/psychotherapy without a mindfulness focus.^26,27,30^ Three were RCTs^26,30^, one of which was a secondary analysis^27^; two studies^26,27^ stated sleep was the primary outcome.

Kiropoulos et al.^30^ compared 8 weeks of CBT to treatment as usual and found group differences in PSQI scores both immediately post intervention and at 20 months follow up. Abbasi et al.^26^ reported significant differences in sleep quality between a group that received 8 sessions of CBT compared to a control group at both the post intervention and one month follow up timepoints. Baron et al.’s study^27^ was a secondary analysis of a study by Mohr et al.^66^ and reported a decline in reported insomnia from 78% as baseline to 43% at post treatment of telephone administered psychotherapy for depression.

### Mindfulness and Relaxation

There were six studies that incorporated mindfulness and/or relaxation.^48,49,58-60,67^ None stated sleep as the primary outcome. One study was an RCT^67^, two were single group pre-post studies^48,49^, one was a quasi-experimental study^60^, and two were qualitative studies consisting of secondary analyses.^58,59^

Cavalera et al’s^28^ RCT reported a strong effect on the Medical Outcomes Study Sleep scale following an eight-week online mindfulness meditation intervention compared to an online psychoeducation control group at the post-intervention evaluation; however, there was no statistical difference between groups after 6 months.

Dayapoglu & Tan’s^48^ six-week single group pre-post progressive muscle relaxation study demonstrated global PSQI score improvement from pre to post intervention and significant improvement in the PSQI subscales of subjective sleep quality, sleep latency, sleep duration, sleep efficiency, sleep disorder, and daytime dysfunction. Hoogerwerf et al.’s^49^ single group pre-post study reported a small non-significant difference between pre, post, and 3-month Symptom Checklist-90 (SCL-90) scores following an eight-week mindfulness-based cognitive therapy intervention.

The intervention group in Lorenz et al.’s^60^ quasi-experimental study showed moderate to large pre-to-post effect on sleep behaviors (not significant), sleep efficiency (significant), and total sleep time (not significant) following an eight-week mindfulness-based stress reduction course study, although both the intervention and control groups had significantly reduced awakenings. There were no statistically significant differences between groups on sleep outcomes at post-intervention, however, there was a small non-significant effect on total sleep time and sleep behavior. Also, a one-way repeated measures ANOVA including baseline, post-intervention, three-month post-intervention for intervention group suggested a non-significant small-to medium effect on self-reported sleep quality and sleep behaviors. Sessanna et al.^59^ was a qualitative secondary analysis of the Lorenz et al. study^60^ and reported that the formats of the sleep retraining program were acceptable and provided an effective mode of delivery to adults with MS. Simpson et al.’s^58^ qualitative secondary analysis of an eight-week mindfulness-based stress reduction study^68^ found that improved sleep was commonly reported.

### Education and Self-management Support

Four studies focused on MS education/fatigue management.^29,36,54,61^ One stated sleep was a primary outcome^36^. Two were RCTs^29^, one of which self-identified as a pilot RCT^36^, one utilized a single group pre-post study design,^54^ and one was a retrospective chart review.^61^

Hugos et al.’s^29^ RCT study compared two 6-week group programs: one was a multi component fatigue management program and the other was a MS education program (control). While there was no significant change in PSQI at program completion or at the 3-month follow-up in the fatigue management group, there was a significant improvement at the six-month follow-up in the fatigue management group compared to the control group; however, PSQI scores remained poor (> 5) at the six-month follow-up timepoint. Akbarfahimi et al.^36^ conducted a pilot RCT comparing an occupational therapy-based sleep intervention and care as usual as a control and found that sleep quality significantly improved in the intervention group compared to the control group.

Sauter et al.^54^ conducted a single group pre-post study assessing the impact of a fatigue management and reported the PSQI global score was significantly improved after the six-week course and remained improved after 7-9 months, and the sleep efficiency component score was also significantly improved at 7-9 months. Participants were initially divided into either a “treatment first” group or a “waitlist” then treatment group; however, all participants were included in the pre-post intervention statistical analyses. The retrospective study^61^ reported significant improvement in sleep disturbances following a 3-4 session individualized cognitive rehabilitation program; however, there was no between groups comparison conducted between the intervention group and the control group.

### Complementary and Alternative Interventions

Five studies utilized complementary or alternative interventions to address sleep issues, including acupuncture,^47^ neurofeedback and mindfulness used in conjunction with hypnosis,^64^ trauma releasing exercises,^52^ foot reflexology,^35^ and integrated imaginative distention.^34^ None included sleep as the primary outcome. Three were RCTs,^34,35^ one of which self-identified as a pilot RCT,^64^ and two utilized a single group pre-post study design.^47,52^

Sajadi et al.’s^69^ RCT reported a significant difference in sleep quality between the group that participated in foot reflexology and the placebo group following the intervention; however, the intervention group’s average PSQI score was significantly worse than the control group’s PSQI score at baseline. Sgoifo et al.’s^34^ RCT reported no significant difference in ISI change scores between the integrative imaginative distention intervention group and the waitlist control group. In Jensen et al.’s pilot RCT,^64^ participants received an intervention with neurofeedback and hypnosis (NF-HYP), mindfulness and hypnosis (MM-HYP), or hypnosis alone (control). There was a large non-significant time x condition effect for sleep disturbance (p=0.17; η2p = 0.15). There was a large significant time effect for the NF-HYP condition which was maintained at the 1-month reassessment, and there was a large non-significant time effect for the MM-HYP condition which was not maintained at the 1-month reassessment.

Lynning et al.’s^52^ study implemented trauma releasing exercise training and found a significant increase in participants sleep quality based on daily self-report measure over the course of the nine-week intervention. Becker et al.^47^ reported a significant improvement in PROMIS sleep disturbance scores from before to after acupuncture and health promotion education.

### Quality Assessment

The details of the quality appraisal analyses are reported in Table 3. The quality score for the twelve RCTs ranged from 35-73%, the seven pilot RCTs ranged from 31-54%, the two quasi-experimental studies were 50% and 88%, and the eleven pre-post single group design studies ranged from 27-64%. The quality score for the two case reports were 50% and 88%, and the two qualitative studies were 70% and 80%. The quality score for the one case series was 81%.

**Table 3.**
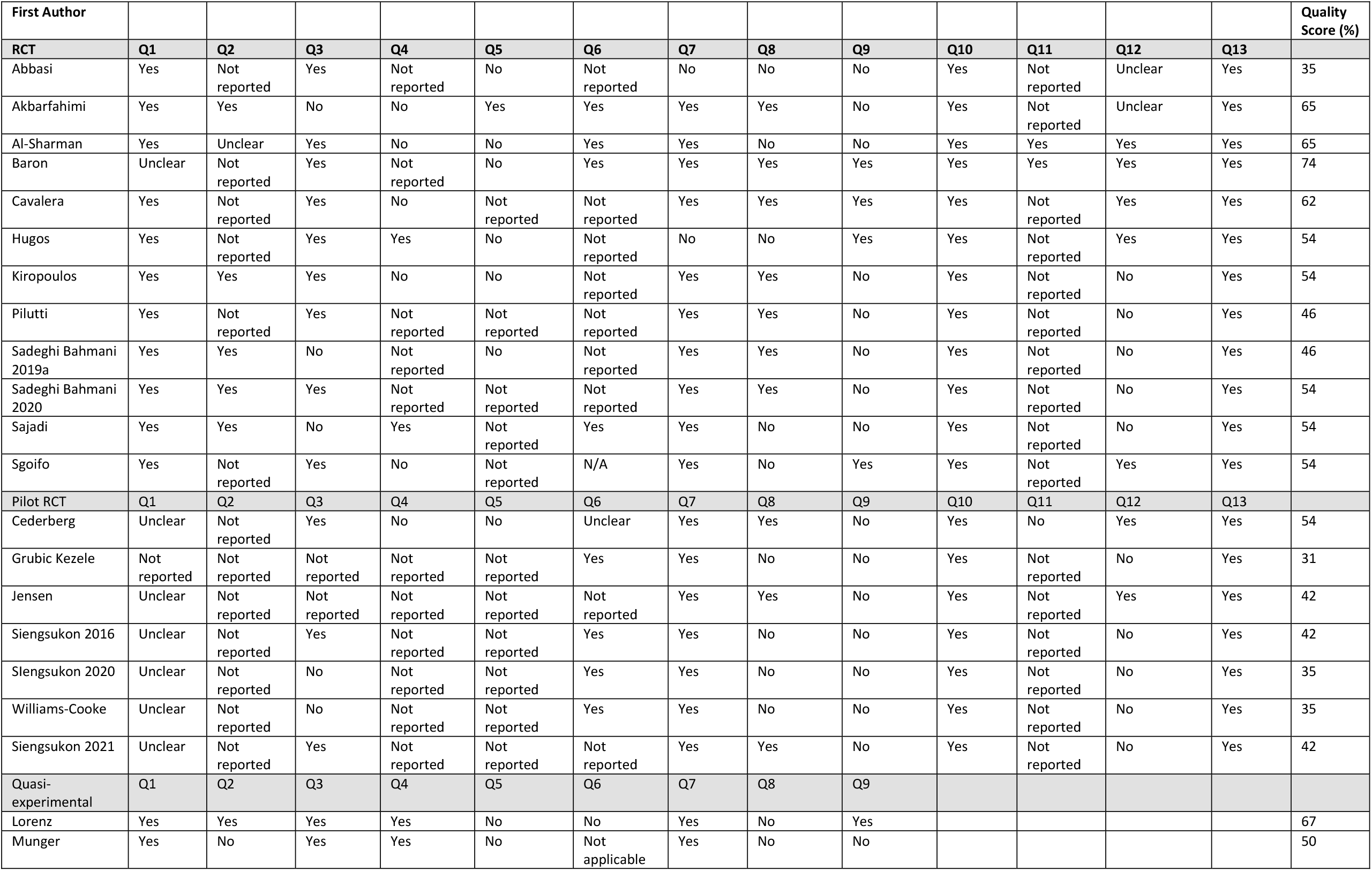
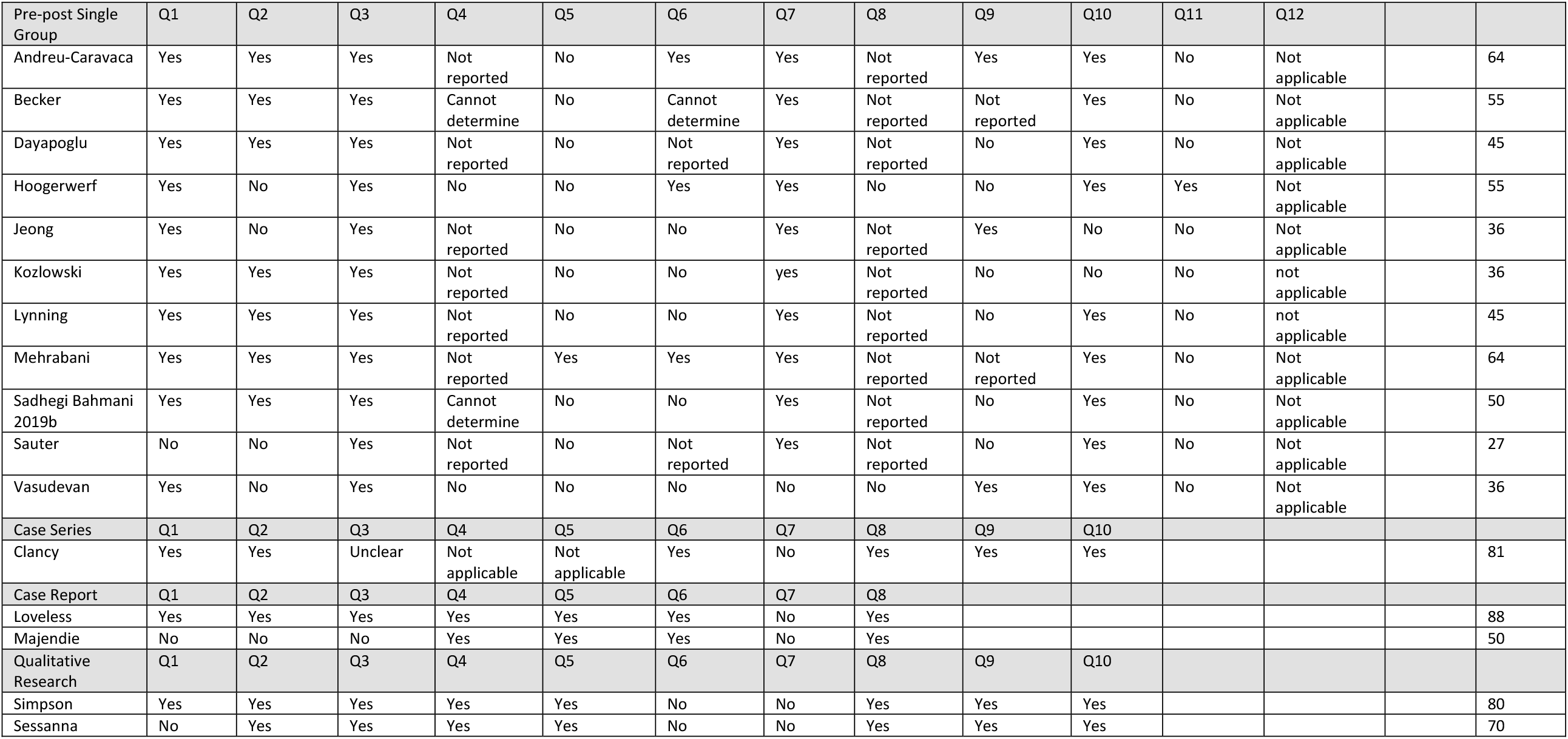
Quality appraisal.

## Discussion

This is the first study to systematically review behavioral interventions to improve sleep in people with MS. The variability in the intervention types, intervention dose, outcomes used, training/expertise of interventionist, specific sample included, and quality of the study made it difficult to compare and synthesize results. Overall, the CBT-I, CBT/psychotherapy, and education/self-management support intervention studies reported positive outcomes, and the physical activity, mindfulness/relaxation, and complementary and alternative intervention studies reported mixed results. The quality appraisal scores ranged from low to high quality indicating potential for bias. While results of this systematic review suggest that many of the interventions are feasible and have a positive impact on sleep in people with MS, further research is necessary to demonstrate efficacy of most of the interventions. More attention to tailoring the intervention to the specific sleep issue, intervention dose, consensus on sleep outcome measures, adequate statistical power, including an appropriate comparison group, ensuring adequate training and experience of interventionists and ensuring intervention fidelity, and appropriate blinding is needed in future research.

Only nine out of 37 studies (24%) explicitly stated sleep was a primary outcome. Depression, fatigue, and feasibility were often named as the primary outcomes. This illustrates the lack of behavioral interventions designed to specifically target sleep in people with MS. What may have contributed to the lack of sleep as a primary outcome is that many of the *interventions* themselves were not designed to target sleep specifically. CBT-I which is a multidimension intervention to specifically treat insomnia was the exception, which likely explains the consensus in improved sleep outcomes in people with MS of the included CBT-I studies. CBT-I, which is a multicomponent treatment strategy that addresses behaviors and thoughts that can negatively impact sleep is the recommended first-line treatment of insomnia.^70,71^ The other interventions included in this review such as mindfulness, relaxation, and complementary and alternative interventions are less likely to target specific sleep impairments or disturbances but instead target secondary contributors to sleep.

It was interesting that the physical activity intervention studies had mixed results (eight had positive results, three had positive results although not statistically significant, and two had minimal to no change in sleep outcomes) due to extensive studies showing physical activity has a positive impact on various sleep outcomes in the general populations.^72-76^ There are several possible explanations for why physical activity had mixed results on sleep outcomes in people with MS including the studies were underpowered to detect meaningful change, the intervention lacked sufficient dose of physical activity to positively affect sleep, sleep impairment was not a requirement for eligibility, or a larger percentage of individuals with more progressive disability were included in the sample. While the exact mechanisms for how physical activity improves sleep are not clear, one possible mechanism is the rise in body temperature due to physical activity leads to self-induced internal cooling processes that initiate sleep.^76,77^ Perhaps due to heat sensitivity in people with MS, the rise in body temperature does not trigger the same internal cooling processes or perhaps other MS symptoms (such as spasticity or pain) mediate the impact on sleep. Additional research is needed to elucidate if physical activity does indeed have a positive impact on sleep outcomes, and if not, then why.

Another challenge with synthesizing the results is the outcomes used to assess sleep were often not valid or reliable measurements of sleep. For example, the primary construct assessed by the NeuroQoL and the Hamilton Depression Rating Scale are not sleep but rather a construct that is associated with sleep; however, these assessments appeared multiple times throughout the review as some studies’ sole sleep outcome measure. Also, interestingly, only six (16%) of the included studies used an objective measure of sleep such as actigraphy or EEG, and no study utilized polysomnography. While objective measures reduce potential bias and assess certain constructs of sleep, they are typically more time consuming and more costly. Perhaps because of the number of preliminary studies or studies with sleep as a secondary outcome or their ease of administration, many studies included in this systematic review utilized self-report sleep assessments. Because objective and self-report sleep measures assess different sleep constructs,^78^ both types of measures should be considered to aid in understanding the effects of behavioral interventions on sleep. Moving forward, studies to assess the impact of behavioral interventions on sleep should use valid and reliable measurements of sleep that are specific to the intended sleep outcome, such as the PSQI to assess self-report sleep quality, the ISI to assess insomnia severity, actigraphy to objectively assess sleep/wake cycle, and polysomnography to assess sleep stages.

Most studies (78%) assessed immediate change in sleep due to the intervention, and only nine studies^26,28-30,40,49,53,54^ included longer-term follow-up (range: one month^26,40^ to seven-nine months^54^). Importantly, three of the studies demonstrated sleep improvements immediately post intervention but were not maintained at long-term follow-up.^28,40,53^ Also, one study did not show positive improvement on sleep at the post-intervention or 3-month follow-up, but did show improvement on sleep at the 6-month follow-up.^29^ This illustrates the importance of including longer-term follow-ups to determine maintenance (or lack thereof) in improvement in sleep outcomes (suggesting maintenance of changed behavior) or that the impact of the intervention on sleep may take time to become apparent. Future studies should include delayed reassessments to determine maintenance of improvement or evolution of change in sleep. In addition, because half of the studies that demonstrated initial improvement in sleep failed to maintain improvement as reassessment, future research should consider emphasizing behavior change theories and techniques.

The quality appraisal that was completed as part of this systematic review highlights limitations and potential biases of the studies included in this review. Many of the studies lacked blinding for participants, interventionists, and/or outcome assessors. Also, many of the RCTs did not include a power analysis and the sample sizes were often small which limits the interpretation of the results. There were numerous participants who dropped out of several studies, and an analysis of the impact of attrition was often absent. These findings from the quality appraisal reveal the potential for bias in the articles included and thus conclusions and interpretation of the results should be viewed with this in mind.

We acknowledge that the JBI critical appraisal checklist for RCTs was used to assess the quality of the self-identified pilot RCTs which likely resulted in a lower quality score for the pilot RCTs. The systematic review team made this decision after thoughtful discussion and after being unable to locate a critical appraisal checklist specifically for pilot RCTs. Pilot RCTs are used to determine feasibility or effect size, while RCTs are used to determine efficacy or effectiveness, and there is debate to whether pilot studies should assess treatment effects or group differences.^79^ There is a need for a critical appraisal checklist specific for pilot RCTs, and the determination of “pilot” should be made *a priori* and in accordance with the purpose and design of the study trial. Another note on the importance of language precision is there was often lack of clarity if the reported MS disease duration was from *diagnosis of MS* or *from symptom onset*. This is an important distinction as it can be years from symptoms onset to actual physician diagnosis of MS.

## Conclusion

Overall, this systematic review has summarized the current state of the literature on behavioral interventions for sleep in individuals with MS and suggests that additional well-designed research with the specific intention of assessing the impact of behavioral interventions on *sleep* using valid and reliable sleep measures in people with MS is warranted. The conclusions in this review and future research identified from this review will aid in guiding clinical practice centered around improving sleep in people with MS.

## Data Availability

All data produced in the present study are available upon reasonable request to the authors

## Acknowledgement

The results of this systematic review will be presented at ACRM 2022

## Abbreviations

CBT: cognitive behavioral therapy
CBT-I: cognitive behavioral therapy for insomnia
ESS: Epworth Sleepiness Scale; International Restless Legs Syndrome Study Group Scale
ISI: Insomnia Severity Index
JBI: Joanna Briggs Institute
MS: multiple sclerosis
NeuroQoL: Quality of Life in Neurological Conditions
PRISMA: Preferred Reporting Items for Systematic Reviews and Meta-Analyses
PROMIS: Patient-Reported Outcomes Measurement and Information System
PROSPERO: Prospective Register of Systematic Reviewers
PSQI: Pittsburg Sleep Quality Index
RCT: randomized controlled trial(s);
RLS: restless legs syndrome
SCL-90: Symptom Checklist-90
WASO: wake after sleep onset

## Notes

### Competing Interest Statement

CS is owner and CEO of Sleep Health Education, LLC

### Funding Statement

This study did not receive any funding

