## Supplementary Tables 1 & 2 for "Behavioral interventions to improve sleep outcomes in people with multiple sclerosis: A systematic review"

**Supplementary File 1--Detailed Search Strategies**

**Embase.com**

(('exercise'/exp OR 'kinesiotherapy'/exp OR 'exercise therap*':ti,ab,kw) OR (cognit* NEAR/3 (therap* OR intervention* OR therap*)):ti,ab,kw OR ('alternative medicine'/exp OR 'mind-body therap*':ti,ab,kw) OR ('psychotherapy'/exp OR 'positive psychotherap*':ti,ab,kw OR 'positive psychology intervention':ti,ab,kw) OR ('muscle relaxation'/exp OR 'progressive muscle relaxation':ti,ab,kw) OR ('qigong'/exp OR qigong:ti,ab,kw) OR ('meditation'/exp OR meditat*:ti,ab,kw) OR ('non pharmacological*':ti,ab,kw OR 'non pharmaceutical*':ti,ab,kw OR 'non drug':ti,ab,kw)) AND (('multiple sclerosis'/exp OR 'multiple sclerosis':ti,ab,kw OR ms:ti,ab,kw) AND ('sleep'/exp OR insomnia*:ti,ab,kw OR sleep*:ti,ab,kw OR asleep:ti,ab,kw OR circadian*:ti,ab,kw))

**Ovid MEDLINE(R) and Epub Ahead of Print, In-Process, In-Data-Review & Other Non-Indexed Citations, Daily and Versions(R) 1946 to December 28, 2021**

1 exp multiple sclerosis/ or multiple sclerosis.ti,ab,kw. or MS.ti,ab. (413212)
2 exp Sleep/ or exp Sleep Wake Disorders/ or (insomnia$ or sleep$ or asleep or circadian$).ti,ab,kw. (291875)
3 exp Exercise/ or exp Exercise Therapy/ or exercis$.ti,ab,kw. or "physical therap$".ti,ab,kw. or physiotherap$.ti,ab,kw. or "physical activit$".ti,ab,kw. (566127)
4 exp Behavior Therapy/ or (cognit$ adj3 (therap$ or intervention$)).ti,ab,kw. (98252)
5 exp Complementary Therapies/ or "mind-body therap$".ti,ab,kw. (242234)
6 exp Psychotherapy/ or positive psycholog$.ti,ab,kw. (210946)
7 Muscle Relaxation/ or "Progressive muscle relaxation".kw,ti,ab. or (muscle adj3 relax$).ti,ab,kw. (27016)
8 (qigong or mindfulness or meditation).ti,ab,tw. (13800)
9 1 and 2 (3171)
10 or/3-8 (994806)

**Web of Science**

#3 AND #4

TI=(exercis* or "behavior therap*" or "complimentary therap* " or "mind-body therap*" or psychotherap* or "positive psycholog*" or relaxation or "progressive muscle relaxation"or qigong or yoga or mindfulness or meditat* or non-pharmacologic* or non-pharmaceutic* or non-drug) OR AB=(exercis* or "behavior therap*" or "complimentary therap* " or "mind-body therap*" or psychotherap* or "positive psycholog*" or relaxation or "progressive muscle relaxation"or qigong or yoga or mindfulness or meditat* or non-pharmacologic* or non-pharmaceutic* or non-drug)

#2 AND #1

Sleep* or insomnia* or sleep* or asleep or circadian* (Title) or Sleep* or insomnia* or sleep* or asleep or circadian* (Abstract)

multiple sclerosis (Title) or multiple sclerosis (Abstract) or MS (All Fields)

| First author Year Country N enrolled (dropouts) | Study design | Study Objective | Sample Demographics | | | | | Intervention/Control Details | | | | Outcomes | | | | Major Sleep Findings |
|---|---|---|---|---|---|---|---|---|---|---|---|---|---|---|---|---|
| | | | Age | MS-type | Severity | Disease duration | Specific impairment(s) for eligibility | Dose | Interventionist | Setting | Control group (if included) | Was sleep the primary outcome | Primary outcome | Sleep outcome | Assessment timepoints | |
| *Physical Activity* | | | | | | | | | | | | | | | | |
| Al-Sharman 2019 Jordan 40 (10) | Pilot RCT | To explore the effects of a six week moderate-intensity aerobic exercise intervention on sleep characteristics and sleep-related biomarkers in people with MS | intervention: 38.7 (13) yo; control: 31.9 (10) yo | Not reported | EDSS intervention: 2.1 (1.8); control: 1.9 (1.03) | intervention: 9.6 (8.49); control: 5.43 (4.2) | >5 on PSQI | 3x/week (approx 50-60 min) for 6 weeks | Not reported | Intervention: supervised setting; control: home | home exercise program 3x/week for 6 weeks | Unclear | Unclear | PSQI, ISI, actigraphy (in subsample), biomarkers | Pre-post | Compared to the HEP, the MAE had significant improvement on the PSQI (p=0.004), ISI (p=0.04), and sleep efficiency (p=0.04) and WASO (p=0.003) assessed via actigraphy; MAE group had significant increase in serotonin compared to the HEP (p=0.002); change in serotonin was significantly correlated with change in PSQI (r = -0.97, p<001 [period missing; unclear if p<0.01 or p<.001]) and ISI (r = -0.56, p = 0.015) for the MAE group. |
| Andreu-Caravaca 2021 Spain 18 (0) | Single group pre/post test | To examine the acute and chronic effects of 10-weeks of progressive resistance training on sleep quality and sleeping heart rate variability in pwMS | 44.88 (10.62) yo | 16 PP, 2 RR | EDSS 3.12 (1.74) | Not reported | mild or moderate disability with mild spastic-ataxic gait disorder | 3x/week for 10 week | supervised by researcher | Sports center | N/A | Yes | actigraphy sleep quality and Karolinska sleep diary | actigraphy sleep quality and Karolinska sleep diary | (1) before the starting of training program on a rest day (RNw1); (2) the night after training week 1 (TNw1); (3) the night after training week 10 (RNw10) and (4) after completin | Sleep quality, sleep comfort, ease of falling asleep, and feeling of rest (from Karolinska Sleep Diary, p<0.05) on the night of no training (RNw1 vs RNw10) improved significantly after 10 weeks of PRT. Actigraphy data showed better sleep efficiency (p = 0.017) on the day they performed PRT session (TNw1) than on the no PRT session (RNw1) at week 1. |

| | | | | | | | | | | | | | | | | |
|---|---|---|---|---|---|---|---|---|---|---|---|---|---|---|---|---|
| | | | | | | | | | | | | | | | g the training program on a rest day (TNw10). | |
| Cederberg 2021 USA 15 (1) | Pilot RCT | To examine the feasibility and efficacy of a physical activity behavior change intervention for improving restless legs syndrome severity and secondary sleep outcomes among a sample of adults with MS | intervention 55.6 (9.5) yo; control 57.4 (13.0) yo | Intervention: 7 (100%) RR; Control: 6 (85.7%) RR, 1 (14.3%) PP | EDSS score median (IQR): intervention 4.0(.5); control 4.0 (1.0) | intervention 22.0(4.4); control 20.0(7.3) | moderate-to-very severe RLS; see inclusion and exclusion criteria | 16 week program with 12 one-on-one video chats | Behavioral coach | home | waitlist control | Unclear | Unclear | International Restless Legs Syndrome Study Group Scale (IRLS), PSQI, Seven-Day Diary, Actigraphy GT3X, ESS | Pre-post | There was a significant and positive effect of the intervention on IRLS scores (F(1,12)=8.43, p=.01, partial eta square=.43). There was a significant and positive effect of the intervention of RLS severity during the night (F(1,12)=6.04, p=.03, pes=.35), during the day while resting (F(1,12)=8.60, p=.01, pes=.44), and during the day while active (F(1,12)=17.35, p<.01, pes=.61). Although not statistically significant, participants reported lower RLS severity while falling asleep than the control group at follow-up (F (1,12) =1.03, p=.33, pes=.09). There was a significant effect of the intervention on sleep satisfaction (F (1,12) =10.68,p=.01, partial eta square =.49), whereby the intervention group reported significantly higher sleep satisfaction compared with the control group following the 16-week period. Although not statistically significant, participants in the intervention group reported lower PSQI scores than the control group following the 16-week period (F(1,12)=0.96, p=.35, pes=.08). Regarding device-measured sleep quality outcomes, there was no significant effect of the intervention on actigraphy-based outcomes. Overall, the intervention group had shorter sleep latency (F[1,11]=1.41, p=.26,ηρ2=.12), longer TIB (F[1,11]=3.77,p=.08,ηρ2=.27), greater frequency of awakenings (F[1,11]=1.75,p=.22,ηρ2=.15), longer total sleep time (F[1,11]=3.29,p=.10,ηρ2=.25), and longer WASO (F[1,11]=1.02, p=.34,ηρ2=.09). There was no effect of the intervention on device-measured sleep efficiency (F[1,11] =0.13,p=.73,ηρ2=.01). Regarding self-reported sleep quality outcomes, there was a significant effect of the intervention on self-reported TIB (F[1,12]=6.47, p=.03,ηρ2=.37) and total sleep time (F[1,12]=6.07,p=.03,ηρ2=.36), whereby the |

| | | | | | | | | | | | | | | | | |
|---|---|---|---|---|---|---|---|---|---|---|---|---|---|---|---|---|
| | | | | | | | | | | | | | | | | intervention group self-reported more time spent in bed and longer sleep duration than the control condition following the 16- week period. Although not significant, the intervention group reported shorter sleep latency (F[1,12]=3.61,p=.08, ηρ2=.25), more awakenings throughout the night (F[1,12]=4.48,p=.058,ηρ2=.29), and better sleep efficiency (F[1,12]=1.36,p=.27,ηρ2=.11) than the control group after the 16-week period. Participants in the intervention group reported lower scores for overall ESS (F [1,12]=0.44,p=.52,ηρ2=.04), RLS-6 daytime sleepiness (F[1,12]= 0.95,p=.35,ηρ2=.08), and average daytime sleepiness based on the Seven-Day Diary (F[1,12]=0.46,p=.51,ηρ2=.04) than the control group after the 16-week period, but the differences were not statistically significant. |
| Grubic-Kezele 2021 Croatia 24 (0) | RCT | To examine the feasibility and possible effect of an 8-week exercise program on sleep quality, insomnia and psychological distress in individuals with multiple sclerosis | intervention: 50.0 (9.3) yo; control: 53.8 (11.8) yo | intervention: 53.8% relapse-remitting, 15.4% primary progressive, 30.8% secondary progressive; control: 36.3% relapse-remitting, 9.1% primary progressive, 54.5% secondary progressive | Intervention (EDSS median (range), mean ± SD): 3.0 (1.0–7.5), 3.8 ± 1.8; control: 5.0 (1.0–7.0), 4.0 ± 2.0 | Not reported | presence of any kind of sleep disturbance determined by the interview | 2x/week, 60 min/session for 8 weeks | physiotherapist | MS Society Center (MSSC) | visit the MSSC for 8 weeks, 2d/week (≤60 min) | Unclear | Unclear | PSQI, ISI | Pre-post | Insomnia severity measured with ISI (F(1;22)=5.95, p = 0.023, ηp2 = 0.213, 90% CI = 0.02–0.42) showed statistically significant group-by-time interaction. Sleep quality measured with the PSQI showed statistically significant group-by-time interaction only in an aspect of daytime sleep dysfunction (F(1;22)=5.33, p = 0.031, ηp2 = 0.195, 90% CI = 0.01–0.40). |

| | | | | | | | | | | | | | | | | |
|---|---|---|---|---|---|---|---|---|---|---|---|---|---|---|---|---|
| Jeong 2020 USA 17 (0) | Single group pre/post test | To investigate the effect of a telerehabilitation system on the quality of sleep in patients with multiple sclerosis (PwMS) | 60.1 (11.4) yo range: 39-76 | Self-report: progressing 5(29.4%), stable 11(64.7%), improving: 1(5.9%) | Self-assessed: Mild 5(29.4%), moderate 10(59.8%), severe 2(11.2%) | 26.0 (13.5), range: 2.5-50.8 (Since diagnosis) | N/A | instructed in individualized exercise plan and asked to perform daily for 3 months (actual average total time over 3mo: 30.7 ± 19.8 hours) | Physical therapist | Home | N/A | Yes | PSQI | PSQI | Pre-post | Average changes between BL and 3M for PSQI sleep efficiency, PSQI sleep disturbances, PSQI daytime disturbances, and PSQI total score were -0.1 (84.7 at BL to 84.5 at 3M), 0.9 (6.4 at BL to 7.3 at 3M), 0.5 (1.0 at BL to 1.5 at 3M) and 0.2 (6.7 at BL to 6.9 at 3M) respectively [indicating a slight worsening in general of sleep from BL to 3M]. The association between PSQI sleep efficiency and Total system usage time and between PSQI sleep efficiency and Total exercise time were 0.757 (p<0.01) and 0.814 (p<0.01) respectively. The association between PSQI total and Total system usage time and between PSQI total and total exercise time were -0.507 (p<0.05) and -0.702 (p<0.01) respectively. |
| Kozlowski 2017 USA 13 (8) | Single group pre/post test | To examine the feasibility, safety, (defined in terms of accessibility, tolerability, risks, learnability, and acceptability) and secondary benefit potential of exoskeleton-assisted walking with one device ('ReWalk Rehabilitation 2.0') for pwMS | median age of 47 yo (range: 38-62) | 92% progressive MS | EDSS 6.5 (5.5-7) | Not reported | walking limitations (EDSS 5-7) | 3x/week 30-90minute sessions for 8 weeks | "Research personnel who completed the device manufacturer's training program" | Outpatient MS clinic/tertiary care hospital | N/A | No | feasibility and safety | Neuro-QoL sleep disturbance section | Weekly throughout BL and the intervention period | 1 had improved sleep disturbance, 1 had worse sleep disturbance, the 6 had no change [note only 5 completed intervention]. |
| Mehrabani 2021 Canada 41 (5) | Single group pre/post test | To examine the efficacy of an intervention focused on sitting les and | 50 (10.3) yo | 26 RR (63.4%) | (EDSS< 4) 18 (43.9%); (4 < /= EDSS</= 6.5) 23 | 14.3 (11.3) | see inclusion and exclusion criteria | 1x/week 15-30 minute sessions for 15 weeks | "Intervention coach" | Home | N/A | Unclear | Unclear | PSQI | Baseline, interim (week 8), immediate post interventi | Significant change from baseline to post intervention for sleep quality (p=.034, effect size .35) but was not sustained from immediate post intervention to long-term follow up. |

| | | | | | | | | | | | | | | | | |
|---|---|---|---|---|---|---|---|---|---|---|---|---|---|---|---|---|
| | | moving more for changing sedentary behavior outcomes, symptoms, QOL, and physical performance in adults with SCI | | | (56.1%) (PDDS= 0) 2 (4.8%); (PDDS< 3) 10 (24.3%); (3</= PDDS</= 6) 29 (70.7%) | | | (two 7 week stages) | | | | | | | on, and 7 weeks post-intervention | |
| Pilutti 2014 USA 82 (6) | RCT | To examine the efficacy of an Internet-delivered, behavioral intervention for improving outcomes of fatigue, depression, anxiety, pain, sleep quality, and HRQOL in 82 ambulatory persons with MS | Intervention: 48.4 (9.1) yo; Control: 49.5 (9.2) yo | RRMS/SPMS/PPMS: Intervention: 31/8/2; Control: 34/2/5 | PDDS, median (IQR) Intervention: 2.0 (4.0); Control: 3.0 (3.0) | Intervention: 10.6yrs (7.1); Control: 13.0 (9.1) (not reported) | N/A | 15 web-based video coaching sessions; seven in the first 2 months, six in the second 2 months, and two in the final 2 months. New content uploaded throughout intervention; daily pedometer log | Behavioral coaches; "highly trained doctoral students or a postdoctoral fellow" | Real-world; web-based video sessions | wait-list control | Unclear | Unclear | PSQI | Pre-post | There was a favorable effect of the intervention on sleep quality post-trial (F[1,73]=3.66, p=.06, np2=.05, Cohen's d = .45), although the differences between groups did not reach statistical significance. |
| Sadeghi Bahmani 2019a Switzerland 51 (5) | Single group pre/post test | The aim of the present study was to investigate the impact of a regular exercise program on subjective and objective | 50.74 (11.28) yo | Not reported; see inclusion/exclusion criteria | EDSS: 5.3; females: M = 5.49, SD = 1.11; males: M = 4.89, SD = 1.27 | not provided | N/A | 5 30 min endurance training sessions per week for 3 weeks; two physiotherapy sessions per day; | Sport scientist; Interventionist for physiotherapy not provided. | Inpatient rehabilitation center | N/A | Yes | Main outcome variables: changes in objective and subjective sleep, depression, fatigue, and | Yes | Pre-post | ISI score was reduced significantly (pre 15.18 (5.71); post 14.00 (4.59); p = 0.048). Sleep onset latency significantly decreased (pre 30.82 (15.63); post 7.81 (8.09);p < .01), sleep efficacy improved (pre 75.39 (14.12); post 82.66 (11.19);p = .04), and wake time after sleep onset diminished (pre 1:57:26 (1:08:59); post 1:22:30 (1:08:59); p = .04). For all other dimensions of sleep continuity (total sleep time, sleep period time, and numbers of awakenings after sleep onset), there were no significant differences between the |

| | | | | | | | | | | | | | | | | |
|---|---|---|---|---|---|---|---|---|---|---|---|---|---|---|---|---|
| | | sleep, depression, paresthesia, fatigue, and cognitive performance in individuals with MS | | | | | | progressive resistance training (45 min) and a low-intensity physiotherapeutic session (30 min) | | | | | paresthesia | | | baseline and the study end. With regard to sleep architecture (stages 1–4 and REM-sleep: min and %), there were no significant differences. |
| Sadeghi Bahmani 2019b Iran 92 (21) females | RCT | To investigate the influence of physical activity on depression, fatigue, sleep, paresthesia, and personality traits (intolerance of uncertainty), and to explore, if endurance training or coordinative training are superior to an active control condition. | Coordination training: 39.17 (8.66) yo; Endurance training: 37.96 (8.69); Active Control: 37.90 (9.91) yo | Not reported | EDDS Coordination training: 3.38 (1.87); Endurance training: 2.46 (1.50); Active Control: 2.02 (1.84) | Groups Coordination: 8.13 (6.37); Workout: 6.92 (6.81); Active Control: 7.21 (6.57) (unclear) | N/A | 8 consecutive weeks 3x/wk 30-45min each | "Professional instructions" monitored intervention; clinical psychologist monitored control | Hospital setting | Active control-30-45min sessions 3x/wk for 8 wks in a hospital setting | Yes | ISI | Yes | Baseline, Week 4, and Week 8 (ISI reported at baseline and week 8) | Sleep complaints decreased over time (time x group interaction, p=<.05, ES=0.32), but more so in the exercise training groups (medium effect sizes), compared to the ACC (small effect size)[p-values not provided for between group comparisons]. Baseline ISI scores for the endurance, coordination, and active control groups were 11.62 (5.23), 13.46 (5.81), and 11.71 (5.43), respectively; 8wk ISI scores for those groups were 8.81 (5.41), 10.13 (4.92), and 11.14 (5.39). Cohen's d for change in ISI from baseline to week 8 for the endurance group, coordination group, and active control were 0.53 (M), 0.62 (M), and 0.11 (S), respectively [p-values not provided for within group change from baseline to week 8]. |
| Sadeghi Bahmani 2020 Iran 62 (2) heterosexual married females | RCT | to examine the effect of aquatic exercise on improving sexual function in females with Multiple Sclerosis | AE 2x/wk: 39.35 (7.10) ; AE 3x/wk: 40.61 (8.97); Active control: 33.77 (6.56) | Not reported | EDSS Median (range) Aquatic exercise (AE) 2x/wk: 3.00 (5.00); AE 3x/wk: 1.5 (4.00); | AE 2x/wk: 8.87 (3.60); AE 3x/wk: 6.58 (3.92); Active control: 6.34 (4.34) (not reported) | N/A | 60 min sessions 2x/wk OR 3x/wk for 8 weeks | "Certified instructor" not otherwise involved in the study organized and supervised | Rehabilitation center of hospital | Active control-60min sessions 2-3x/wk for 8 wks in a hospital setting | No | Sexual dysfunction | No | Pre-post | There was an non-significant intervention effect on sleep (p=0.079, np2=0.09) "whereby the 2x/w exercise condition had better scores than the other groups (medium effect size). Additionally, sleep complaints were lower in the 3x/w exercise condition than ACC, although the difference was not statistically significant." [omnibus post-trial ISI ANCOVA was not identified as statistically significant; no p-values given for between group comparisons; sleep complain data provided in "pre-trial" Table 1 is same as in "post-trial" |

| | | | | | | | | | | | | | | | | |
|---|---|---|---|---|---|---|---|---|---|---|---|---|---|---|---|---|
| | | | | | Active control: 1.50 (6.00) | | | | aerobic exercise; Control: clinical psychologist | | | | | | | Table 2 which makes interpretation of results difficult] . |
| Siengsukon 2016 USA 33 (3) | RCT | to examine the efficacy of a supervised moderate-intensity aerobic exercise program and an unsupervised low-intensity walking + stretching program on sleep quality and daytime sleepiness in PwMS | Intervention:48.9 (13.6) yo; Control: 50.9 (12.2) yo | 19 RR, 3 SP (10 RR, 2 SP in intervention analysis, 9 RR, 1 SP in control) | MSFC 0.395 (0.89) and 0.868 (0.68) | 10.8 ( 8.4), 9 (5.6) (not reported) | N/A | 3x/week hour long sessions for 12 weeks | Not reported | Not reported | active control low impact walking and stretching | No | feasibility | ISI, PSQI, and SSE | Pre-post | Both groups improved on PSQI (main effect sleep p=0.006; np2=0.336) but no sig between group difference (p=0.447; np2=0.031); there was however a significant between group difference on ESS (p=0.018; np2=0.263): AE group had a 28.6% reduction (p=0.016) and WS had a 7.9% increase (p=0.447) in daytime sleepiness. |
| Vasudevan 2021 India 10 (0) | Single group pre/post test | to assess the feasibility of a personalized yoga therapy intervention on quality of life, sleep quality, and symptom relief among patients with MS | Not reported, see inclusion/ exclusion criteria | Not reported | Not reported | First MS attack occurred: n =3 <10 years ago, n=5 11-19 years ago, n=2 20+ years ago | n=2 poor sleep; other physical/emotional issues reported | 12 private (1 therapist: 2 patient ratio) hour-long weekly sessions over 3 months and 3 group sessions of education, sessions lasted 90-100 minutes | IAYT-certified yoga therapists | Private setting | N/A | Unclear | Unclear | PSQI | Pre-post | Non-significant (p=0.061) improvement in sleep quality; decrease from median of 8 (5-15) to 6 (5-11); "sleep" listed as an improved outcome "reported by patients and observed by therapists" [unclear how patient report and therapist report determined]. |
| *CBT-I* | | | | | | | | | | | | | | | | |

| | | | | | | | | | | | | | | | | |
|---|---|---|---|---|---|---|---|---|---|---|---|---|---|---|---|---|
| Clancy 2015 USA 11 (unclear) | Case series | To examine outcomes of cogntive behavioral therapy for insomnia (CBT-I) in individuals with MS | 52(11); range: 36-69 | 8 RR; 3 primary or secondary progressive | Not reported | Not reported | insomnia | between 2 to 16 sessions (mean: 8 (4.8)) | "Clinician" | Not reported | N/A | Unclear | Unclear | ISI, TST | Unclear | Most individuals (86%) reported improvement regarding their insomnia with an average ISI score 21 before CBT-I and average ISI score of 17 after treatment (ISI scores recorded for only 7 of 11 participants because ISI not used by clinic until 2010. Most individuals (73%) reported an increase in TST, with an average improvement of 1.5 hours. Before CBTI, the average reported TST was 5.5 hours compared with 7 hours after treatment. The TST was recorded for all the participants at the pretreatment and posttreatment intervals. |
| Loveless 2019 USA 1 (n/a) male | Case study | To provide practical guidance to practitioners regarding modifications made to the CBT-I protocol for an individual with MS | 55 | RR | Not reported | started in early 1990s | Multiple; self-report and medical record | biweekly, 13 sessions in total | postdoctoral fellow and a practicum student supervised by an attending licensed clinical psychologist | University health system behavioral medicine clinic | N/A | Unclear | Unclear | Self-report sleep quality, sleep log, ISI, and ESS | start of intervention, 2 week sleep diaries toward beginning of care, sleep diary reviewed at each session; final session | Patient reported improvement in self-report sleep quality; sleep log demonstrated an increase in sleep efficiency (from 29.0% to 74.49%) and TST (from 2.69 hrs to 5.86 hrs) and decrease in sleep latency (from 145 min to 60 min), WASO (from 240 min to 60 min), and early morning awakenings (from 110 min to 0 min); At post-treatment, scored 6/28 on ISI and 2/24 on Epworth Sleepiness Scale. |
| Majendie 2017 UK 1 (n/a) male | Case study | To share case study on an individual with MS who had been experiencing sleep problems and had reported dependency on sleep medication, who was treated with a CBT-I program over the course of 3 months, | 40 male | SP | wheel chair bound' (but no measure explicitly reported) | At least 9 years | Sleep disturbance | 1x/week, 1hr sessions for 8 weeks, plus homework with each session | psychologist | Local Community Neuro Service Clinic | N/A | Unclear | Unclear | PSQI and sleep diary | Assessment of sleep problem took place over two sessions but was also ongoing throughout the treatment period | Improvement in global PSQI from 10 at start to 5 at end of treatment and to 1 at 7 months post treatment; overall improvements in sleep log outcomes. |

| | | | | | | | | | | | | | | | | |
|---|---|---|---|---|---|---|---|---|---|---|---|---|---|---|---|---|
| | | with the goal of improving sleep quality while reducing sleep medication | | | | | | | | | | | | | | |
| Siengsukon 2021 USA 41 (21) | Pilot RCT | To assess the feasibility (primary purpose) and treatment effect (secondary purpose) of CBT-I delivered using web based application with or without biweekly telephone calls to improve sleep quality and fatigue in individuals with MS and symptoms of insomnia | Intervention: 50.1 (11.8); Control: 53.8 (6.9) | wCBTi: 10 RR; wCBTi + calls 9 RR, 1 SP | PDDS wCBTi: 1.3 (1.5) and wCBTi + calls: 2.4 (1.3) | Not reported | insomnia | 41 total daily lessons over 6 weeks (with or without one-on-one telephone calls) | trained in motivational interviewing | Not reported | Webbased CBT-I without telephone calls | No | feasibility | ISI, PSQI, SSES, and MCSB | Pre-post | Both groups demonstrated significant improvement in ISI (wCBT-I: p<001; wCBT-I + calls: p=0.005) but no significant difference between groups (p=0.068); eight of the ten participants in the wCBT-I group and five in the wCBT-I + calls group showed an MCID of at least 6 points on the ISI; both groups showed significant improvements in PSQI (wCBT-I: p<001; wCBT-I + calls: p=0.028). Seven of the ten participants in the wCBT-I group and four of ten in the wCBT-I + calls group showed an MCID of 3 points or more on the PSQI. |
| Siengsukon 2020 USA 33 (3) | RCT | To assess the feasibility (primary aim) and treatment effect (secondary aim) of using CBT-I to improve MS symptoms of reduced sleep quality and fatigue in PwMS with | CBTi: 51.1 (7.9), AC: 50.4 (12.4), BE: 56.9 (10.1) | RR/SP: CBTi 8/2, AC 10/0, BE 9/1 | PDDS CBTi 1.3 (2.21), AC 1.7 (2.3), BE 2.0 (2.1) | CBTi 17.3 (8.5), AC 9.1 (8.9), BE 18.3 (11.4) (Not reported) | insomnia | 1x/week, 45-60 min one-on-one for 6 weeks | trained CBT-I specialist/PT | Not reported | 3 arms in this study (CBT-I, active control or 1-off breif education) Active control did stretching and self-selected | No | feasibility | ISI, PSQI, and SSES | Pre-post | All 3 groups showed a significant large reduction on ISI (CBT-I: p<0.001, ES=2.729; AC: p=0.002, ES=1.018; BE: p<0.001; ES=2.570) with the CBT-I showing the largest change (-13.8); CBT-I and BE showed a significant large improvement on PSQI (CBT-I: p<0.001, ES2.314; BE: p<0.001, 1.672) with CBT-I having largest magnitude in change (-6.7); CBT-I and BE group showed significant increase in SSE (CBT-I: p<0.001; BE: p=0.044) but only CBT-I showed a large effect (ES=1.221). |

| | | | | | | | | | | | | | | | | |
|---|---|---|---|---|---|---|---|---|---|---|---|---|---|---|---|---|
| | | symptoms on insomnia | | | | | | | | | light or sedentary activities in person same dosage as intervention | | | | | |
| Williams-Cooke 2021 USA 25 (3) | Pilot RCT (secondary analysis of Siengsukon 2020) | This study aimed to determine if CBT-I improves sleep log and actigraphy outcomes in individuals with MS. | CBTi: 49.25 (7.72) yo, AC: 50.38 (13.96), BE: 58.78 (8.66) | RR/SP CBTi 7/1, AC 8/0, BE 8/1 | Not reported | CBTi 13.88 (5.14), AC 8.88 (10.02), BE 19.22 (11.72) (Not reported) | Insomnia | 1x/week, 45-60 min one-on-one for 6 weeks | trained CBT-I specialist/PT | Not reported | 3 arms in this study (CBT-I, active control or 1-off breif education) Active control did stretching and self-selected light or sedentary activities in person same dosage as intervention | Yes | Sleep | Actigraphy and sleep log data | Pre-post | For the sleep log outcomes, the CBT-I group showed a significant increase in SE (p = 0.006, ES = 1.203) and reduction in TIB (p = 0.001, ES = 0.993) and variability in SE (p = 0.026, ES = 0.657) from baseline to post-intervention, but no change in the other sleep log variables. The EC and AC groups did not show a significant change from baseline to post intervention for any of the sleep log outcomes. For the actigraphy outcomes, the CBT-I group demonstrated a significant reduction in TIB (p = 0.005, ES = 0.925) and TST (p = 0.004, ES = 0.893) from baseline to post-intervention. The active control group showed a significant increase in variability in SE (p = 0.035, ES = 0.463). There were no significant changes for the other actigraphy variables. |
| *CBT/Psychotherapy* | | | | | | | | | | | | | | | | |
| Abbasi 2016 Iran 72 (6) females | RCT | To evaluate the effect of group cognitive behavioral treatment on the quality of sleep in women with MS | intervention: 85.2% relapsing-remitting; control: 88.3% relapsing-remitting | Not reported, see inclusion/exclusion criteria | intervention: 5.6 (5.8); control: 6.1 (6.5) | 8 90-min sessions, 1x/week, group | ≥5 on PSQI | psychiatric nurse (the researcher) | Not reported | intervention: 35.3 (5.3); control 33.2 (8.9) | 3 group sessions to talk about their diease and experiences | Yes | PSQI | PSQI | pre-post and one month after intervention | Between group: significant differences were seen between the mean score of sleep quality for the control vs. intervention groups at post intervention and one month follow-up (p<0.001). within group: rmANOVA showed that the mean score of sleep quality of patients in the intervention group had a significant difference (p<0.001) across the three time points (baseline, post, follow up) but there was no significant change over time for the control group. |

| | | | | | | | | | | | | | | | | |
|---|---|---|---|---|---|---|---|---|---|---|---|---|---|---|---|---|
| Baron 2011 USA 127 (7) | RCT (secondary analysis) | To determine the prevalence of insomnia in patients with multiple sclerosis and depression, to evaluate the pretreatment correlation between insomnia, anxiety and MS symptoms, to determine the effects of treatment for depression on insomnia symptoms, and to determine treatment-related factors associated with improvement in insomnia, including improvement in depression and anxiety symptoms as well as treatment type | 89% RR, PP 10%, 1 did not report type | Guy's Neurological Disability Scale: CBT mean 23.9, SD 5.8; SEFT mean 22.9, SD 6.7 | Not reported | 16 weekly sessions, 50 min each | ≥ 16 on Beck Depression Inventory-II and ≥ 14 on the Hamilton Depression Rating Scale | doctoral level psychologists with 1-5 years of post degree experience | Telephone | CBT: 48.9 (9.6); SEFT: 47.6 (10.1) | psychotherapy - nonspecific therapy effects | Yes | 3 items on the Hamilton Depression Rating Scale | 3 items on Hamilton Depression Rating Scale | baseline, mid treatment (week 8), and post treatment (week 16) | 78% (n=98) reported insomnia of any type 3 or more times/week at baseline which declined to 43% (n=53) at post treatment [did not report within group change in insomnia or between group comparison in Results]. |
| Kiropoulos 2016 Australia 30 (0) | RCT | To examine the effectiveness and acceptability of an 8-week individual tailored | RR | 100% "Able to walk independently without aid" | Since diagnosis /since first symptoms (months): | 1x/week 1hour sessions for 8 weeks (first session 1.5hr) | ≥ 10 BDI-II | A senior clinical psychologist and a provisional clinical psychologist | Outpatient hospital clinic | CBT: 34.60 (9.06); TAU: 39.27 (9.93) | treatment as usual | No | Depression | PSQI | pre-post and 20wk follow-up | There were significant group differences on the PSQI score post intervention (p<0.001) and at 20 week follow-up (p<0.01). Pre/post/20wk PSQI scores for the CBT group were 9.00(3.46)/4.40(2.09)/4.80(2.54). Pre/post/20wk PSQI scores for the TAU group were 9.06(3.93)/7.87(2.99)/8.20(3.60). |

| | | | | | | | | | | | | | | | | |
|---|---|---|---|---|---|---|---|---|---|---|---|---|---|---|---|---|
| | | cognitive behavioural therapy (CBT) intervention for the treatment of depressive symptoms in those newly diagnosed with multiple sclerosis. | | | Intervention: 26.20 (15.58)/ 35.54 (16.47) Control: 23.53 (16.06)/ 30.57 (18.68) | | | | | | | | | | | Cohen's d for PSQI pre to post was 1.31 and pre to 20mo follow up was 1.06. |
| *Mindfulness/Relaxation* | | | | | | | | | | | | | | | | |
| Dayapoglu 2012 Turkey 35 (3) | Single group pre/post test | To investigate the effect of progressive muscle relaxation technique (PMRT) on fatigue and sleep quality in patients with MS | 38.15 (9.48) | Not reported | Not reported | < 2 yr 43.8%, 3-5 years 18.8%; 6-8 yrs 15.5%; >8 yrs 21.9% (not reported) | ≥ 4 on Fatigue Severity Scale | 1 hour long patient education session about PMRT under supervision of instructor; After education, perform the exercises as home 1x/day for 6 weeks; Two weeks after education session, participants came to clinic and performed PMRT under supervisi | Researcher; no details provided on who provides education | Neurology polyclinic; home | N/A | Unclear | Unclear | PSQI | Pre-post | Global PSQI score decreased from before PMRT (10.81, SD 4.01) to after PMRT (6.25, SD 3.34, p<0.001); There was a significant improvement in the PSQI subscales of subjective sleep quality (p<0.001), sleep latency (p<0.001), sleep duration (p<0.05), sleep efficiency (p<0.05), sleep disorder (p<0.001), and daytime dysfunction (p<0.001). |

| | | | | | | | | | | | | | | | | |
|---|---|---|---|---|---|---|---|---|---|---|---|---|---|---|---|---|
| | | | | | | | | on of the instructor | | | | | | | | |
| Hoogerwerf 2017 Netherlands 59 (20) | Pilot RCT | To study the feasibility and potential effectiveness of mindfulness-based cognitive therapy in severely fatigued multiple sclerosis patients | 48.2 (8.5), range: 32–60 | 67% RR, 33 SP | EDSS 3.9 (1.7) [range=1–7.5] | 11.2 (7.9) [range:1–32] (not reported) | ≥35 on CIS-20-Fatigue | 8 weekly group meetings of 2.5 h (20 h in total) spread over a period of 10 weeks, with homework and exercises of up to 1 h, for 6 days a week. | certified MBCT trainers with a minimum of 2 years of experience with MBCT and in working with MS patients | Not reported | waiting list within-subjects control - standard care | No | CIS-20-Fatigue | sleep subscale of the SCL-90 | pre-waitlist, post-waitlist, post treatment, and 3 month follow up | For SCL-90-sleep, the effect size was small (partial η2= 0.08). Based on an alpha value set at 0.01, the change in SCL-90-sleep was not significant at the different assessment time points(p=0.030). |
| Simpson 2018 Scotland 33 (0) | Qualitative (secondary analysis) | To gather views from two groups of participants with MS after completing an 8-week MBSR course. The first group completed the 'standardized' course and their input was used to inform and test an optimized course which the second group completed | range: 21-66 | Mixed, but mostly relapse remitting | EDSS range 1-7 (mean 4.5) | Not reported | N/A | 1x/week for 8 weeks | MBSR instructor | NHS center for integrated care | N/A | No | Unclear | Qualitative reports | post intervention | Improved sleep was commonly reported |
| Cavalera 2019 Italy 139 (49) | RCT | To test the efficacy of an online mindfulness- | intervention: 42.26 (8.35); control: | intervention: 94% RR, 6% SP; | EDSS: intervention median | Since MS diagnosis: intervent | N/A | 1x/week group sessions | expert MBSR trainer | Skype | psychoeducation | No | Multiple Sclerosis Quality of Life-54 | Medical Outcomes Study Sleep scale | baseline, post-intervention, 6 | A strong effect of the mindfulness program was found on sleep at the post-intervention evaluation (p<0.001, np2=0.130), but no |

| | | | | | | | | | | | | | | | | |
|---|---|---|---|---|---|---|---|---|---|---|---|---|---|---|---|---|
| | | based intervention (MBI) to improve quality of life, psychological well-being, sleep, and fatigue | 43.19 (9.02) | control: 92% RR, 8% SP | 3; control median 3 | ion mean 12.64 (8.15); control mean 14.24 (7.27) | | for 8 weeks | | | | | | | months post-intervention | statistical difference between groups was found after 6 months (p=0.202, np2=0.017). |
| Lorenz 2021 USA 34 (16) | Quasi-experimental | To explore the feasibility of 'SleepWell!' (MSBR plus sleep education) delivered via videoconference or in-person; secondary - to examine preliminary efficacy on sleep and impact on common MS symptoms (depressive, fatigue, function, QoL), sleep promoting behaviours and mindfulness | 47.1 (10.9) | Not reported | EDSS 1.6 (1.1) | Since diagnosis: 11.2 (6.5); since symptom onset: 14.5 (9.0) | N/A | 1 orientation session followed by 1hr/week training for 8 weeks | certified MBSR instructor and a 'sleep expert' (researchers from nursing) | mainly virtual but in clinic comparator | waitlist control (active group had a virtual group and an onsite group) | No | feasibility of intervention | Actigraphy, PSQI, The Functional Outcomes of Sleep Questionnaire-10 (FOSQ-10), and The Sleep Behavior Self-Rating Scale-Revised | pre-post and 3 months | Control and Intervention sleep efficiency scores, respectively: (pre (SD) post (SD), p): 62.4 (14.1) 57.3 (17.1) 0.588; 49.2 (9.9) 60.9 (18.8) 0.042. Pre- Post-intervention within groups: Paired t-tests showed that the videoconference and control groups had reduced number of awakenings suggesting results were independent of any study effects (p=0.039 and p=0.032, respectively). In the videoconference group, Cohen's d effect sizes suggested a moderate to large pre to post effect on sleep behaviors (p=0.145; d=0.36), sleep efficiency (p=0.042; d=0.78), and total sleep time (p=0.156; d=0.54). Post-intervention between groups: Independent t-tests showed no statistically significant differences between groups in sleep outcomes. Cohen's d suggested a small effect on total sleep time (p=0.522; d=0.35) and sleep behavior (p=0.458; d=0.33). One-way repeated measures anova including baseline, post-intervention, three-month post-intervention for videoconference group: Partial eta-squared suggested a small-to medium effect on self-reported sleep quality (p=0.415; partial η2=0.278) and sleep behaviors (p=0.069; partial η2=0.257). |
| Sessanna 2021 USA 14 (0) | Qualitative (secondary analysis of Lorenz 2021) | The purpose of this qualitative descriptive study was to explore the experience of participating in an online or traditional onsite 8- | Not reported; between 30 and 70 | Not reported | Not reported; EDSS < 7 | Not reported | N/A | 1 orientation session followed by 1hr/week training for 8 weeks | certified MBSR instructor and sleep expert | Mainly virtual but inclinic comparator | N/A | No | acceptability | Qualitative reports | 1 time focus group (post-intervention) | Both formats proved to be acceptable and provide an effective mode of delivery to a variety of adults with MS who experience a wide variety of symptoms and levels of disability. |

| | | | | | | | | | | | | | | | | |
|---|---|---|---|---|---|---|---|---|---|---|---|---|---|---|---|---|
| | | week, once a week, Mind Body Stress Reduction combined with Sleep Retraining (SleepWell!) course among women living with MS to establish online course acceptability. | | | | | | | | | | | | | | |
| *Education and Self-management Support* | | | | | | | | | | | | | | | | |
| Akbarfahimi 2020 Iran 22 (2) | Pilot RCT | To design a sleep-targeted intervention program for MS and examine the efficacy of the intervention compared to traditional occupational therapy on sleep quality (primary outcome), fatigue and quality of life (secondary outcomes) | intervention: 37.5 (8.89); control: 39.7 (7.90) | Not reported | EDSS intervention: 1.6 (0.97); control: 1.2 (1.03) | not reported | ≥5 on PSQI | 8 30-45 min sesssions, 1x/week + 2-3 sessions/ week by telephone | occupational therapist | Occupational therapy clinic | Care As Usual 8 sessions 30-45 min, 1x/week | Yes | PSQI | PSQI | Pre-post | Sleep quality for the intervention group significantly improved from baseline (15.2 (2.44)) to reassessment ((mean ± SD = 7.2 ± 1.03; p<0.001). No statistically significant improvement was seen in the control group (baseline 12.8 (2.85) to reassessment 13.2 ± 3.48). In the intervention group, sleep quality improved significantly across all items (p<0.001, effect size = 0.60) except for sleep efficiency and the use of sleep medications. |
| Hugos 2019 USA 218 (14) | RCT | To determine whether *Fatigue: Take Control* (FTC) program is associated with greater improvements in fatigue than *MS: Take Control* (MSTC), a | Intervention (FTC): 53.9 (9.8), range: 28-78; Control (MSTC): 53.6 (10.5), range: 28-73 | (RRMS, SPMS, PPMS), % Intervention (FTC): 62.6, 14.0, 24.3; Control (MSTC): | EDSS mean (SD, range) Intervention (FTC): 5.1 (1.1, 3.5-6.5); Control (MSTC): | Intervention (FTC): 12.3 (7.6) range: 1.59-34.96; Control (MSTC): 12.7, (9.3) range: 0.25- | ⩾25 on the MFIS | 1x/week 2 hour group sessions for 6 weeks | MS professionals | Outpatient MS clinic | Multiple Sclerosis: Take Control (MSTC) group education program | No | MFIS | PSQI | Baseline, immediate post intervention, 3 month follow up, and 6 month follow up | There was no significant change in PSQI either at program completion (p=0.09) or at the 3-month follow-up in the fatigue education group (p=0.83). Sleep quality improved in the FTC group at 6 months (mean change −1.14 points (p < 0.0001) and there was a significant difference between groups by this time point (p = 0.004; Table 2); However, sleep quality (PSQI scores) improved at 6 months in the FTC group compared to the MSTC group but remained poor (PSQI > 5). |

| | | | | | | | | | | | | | | | | |
|---|---|---|---|---|---|---|---|---|---|---|---|---|---|---|---|---|
| | | similarly structured general MS education program | | 55.6, 19.4, 24.1 | 5.3 (1.1, 3.0-6.5) | 41.78 (from diagnosis ) | | | | | | | | | | |
| Munger 2021 USA 21 (0) | Quasi-experim ental - retrospe ctive chart review | To characterize changes in quality of life measures associated with participation in a clinical individualized cognitive rehab program for pwMS | Interventi on: 75% RR; comparat or: 67% RR | Not reporte d | Not reported | varied, individual ized, on average 3-4 sessions | Cognitive | Occupati onal therapist and SLP | Real-world clinic | interven tion 47.9 (4); compara tor 48.9 (4.4) | Compara tor group of individual s who qualified for the cog rehab training program but didn't attend | No | NeuroQoL | Sleep disturbance domain on QoL measure | Pre-post | Sleep Disturbance scores improved after participation in the ICRP (50.5 from 55.5, p = 0.005); no significant change in Sleep Disturbance in the comparison group. [no between group difference provided]. |
| Sauter 2008 Austria 32 (5) | Single group pre/post test | To investigate long term effect of a fatigue management course | Not reported | 53% RR, 47% progres sive (13.3% primary , 33.3% seconda ry) | EDSS 4.0 (1.9) | 11.8 (7.2); range 2.5-27.3 | reported suffering from fatigue for at least 6 months; fatigue had to have a significant impact on patients' life | 1x/week group sessions for 6 weeks, 2 hour sessions | Not reported | Not reported | waiting control group but appears collapsed into 1 group for analysis | Unclear | not stated | PSQI | baseline, immediat ely after participati on in the course and 7-9 month follow-up | The PSQI was significantly improved after the course (from 7.9, SD 4 to 5.8, SD 2.3) and remained improved after 7-9 months (5.9, SD 3.5; p=0.004); the sleep efficiency component score was significantly reduced at 7-9 months (p=0.047). |
| *Complementary and Alternative Interventions* | | | | | | | | | | | | | | | | |
| Becker 2016 USA 14 (0) | Single group pre/post test | To explore change over time in symptom management, health promotion, and quality of life following exposure to a holistic intervention combining group | 1 Benign sensory, 6 RR, 2 PP, 2 SP, 3 unclear | Not reporte d | 13.64 (7.99) (diagnosi s) | 8 90-minute classes with acupunct ure before or after each 30 minute class | N/A | nurse certified in MS nursing // delivered by the third author, who is a licensed and national board–certified | Not reported | 54.14 (9.65) | N/A | Unclear | Not stated | PROMIS sleep disturbance | Pre-post | Pre to post PROMIS sleep disturbance scores were 22.93 (7.82) and 17.21 (3.31) (p=0.009), respectively. This was validated by the women's comments in the focus groups about their perceptions of the effect of the intervention improving sleep. |

| | | | | | | | | | | | | | | | | |
|---|---|---|---|---|---|---|---|---|---|---|---|---|---|---|---|---|
| | | acupuncture with group sessions about health promotion for women with multiple sclerosis | | | | | | acupuncturist, or one of her clinical colleagues | | | | | | | | |
| Jensen 2018 USA 35 (3) | pilot RCT | To determine if neurofeedback and mindfulness meditation training enhances the efficacy of hypnosis for decreasing pain and fatigue in pwMS. | 54.14 (9.65) | 1 Benign sensory, 6 RR, 2 PP, 2 SP, 3 unclear | Not reported | 13.64 (7.99) (diagnosis) | chronic pain, chronic fatigue, or both | 8 90-minute classes with acupuncture before or after each 30 minute class | nurse certified in MS nursing // delivered by the third author, who is a licensed and national board–certified acupuncturist, or one of her clinical colleagues | Not reported | Hypnosis Intervention | No | Average pain intensity/fatigue severity | PROMIS Sleep Disturbance Short Form 8-item Version B | Baseline (pre NF, MM, or Wait), prehypnosis, posthypnosis, and 1mo follow-up | A large interaction effect size (η2p = .15) emerged for sleep disturbance (not significant p=.17). There was a large significant time effect for the NF-HYP condition (p=0.03; η2p = 0.61) and a large non-significant time effect for the MM-HYP condition (p=0.40; η2p = 0.33). Participants in the NF-HYP condition reported average baseline PROMIS Sleep Disturbance scores that were more than .50 of a standard deviation unit above the national norms (mean = 55.64), and these improved to normative values after intervention (mean = 48.62); this improvement for the NF-HYP participants was maintained at follow-up (mean = 50.23). Participants in the MM-HYP condition also evidenced meaningful improvements in sleep disturbance from baseline (mean = 54.85) to posthypnosis (mean = 52.88). However, their scores returned to their baseline levels at follow-up (55.92). |
| Lynning 2021 Denmark 9 (2) | Single group pre/post test | To test a Trauma Releasing Exercise program for pwMS and explore outcome measures to be applied in future randomized trials | 57.53 (10.63) | MS onset course: 23 (72%) RR, 8 (25%) Progressive, 1 (3%) Not reported; Current disease course: 17 | Not reported | Since MS diagnosis: 20.09 (10.00); Since MS symptoms began: 28.52 (13.63) | see inclusion and exclusion criteria | 6 sessions (3 weeks 2x/week) of either (a) neurofeedback or (b) mindfulness meditation followed by a single face-to- | MM-HYP: 1 of 3 bachelor-level research staff who were trained and supervised by a clinical psychologist expert in | Outpatient Medical Center or MS clinic | N/A | Unclear | Unclear | 5-point numeric rating scale in newly developed patient reported outcome tool | Weekly | Sleep quality significantly increased (p<0.05) based on self-reported sleep quality on a 5-point Likert scale. |

| | | | | | | | | | | | | | | | | |
|---|---|---|---|---|---|---|---|---|---|---|---|---|---|---|---|---|
| | | | | (53%) RR, 6 (19%) SP, 3 (9%) PP, 6 (19%) uncertain | | | | face hypnosis session and then four sessions of neurofeedback/mindfulness meditation provided just before four (audiotaped) self-hypnosis training sessions or (c) 3 weeks 2x/week of a waiting period followed by a single face-to-face hypnosis session and then four audiotaped self-hypnosis training sessions | mindfulness-based treatments; NF: staff; in-person HYP: clinical psychologist experienced with hypnosis | | | | | | | |
| Sajadi 2020 Iran 70 (7) | RCT | To indicate the role of reflexology as a complementary therapy on | 51.6 | 6 RR, 1 SP, 2 PP | Not reported | 12.7 (since diagnosis) | N/A | weekly for 9 weeks with instructor and daily | TRE instructor + home | home and not provided info about sessions | sham/non-specific foot reflexology | Unclear | Unclear | PSQI | Pre-post | There was a significant difference between the two groups' PSQI scores at baseline (sleep quality of the intervention group is poorer than the control group; 8.39 ± 2.78 & 6.73 ± 3.02; p=0.04). The difference in sleep quality between the reflexology and control groups |

| | | | | | | | | | | | | | | | | |
|---|---|---|---|---|---|---|---|---|---|---|---|---|---|---|---|---|
| | | the improvement of fatigue, sleep quality, and anxiety in MS patients | | | | | | at home after the second session (session were between 30 and 45 min | | with instructo r | | | | | | was statistically significant after the intervention (5.76 ± 2.56 and 10.03 ± 7.96; P = 0.001). Also, in the reflexology group, the difference in PSQI scores before and after the intervention was statistically improved (P = 0.003). |
| Sgoifo 2017 Italy 48 (0) | RCT | To evaluate the effectiveness of Integrated Imaginative Distention therapy on fatigue in three chronic, stress-related conditions (including MS) | range: 18-50 | 100% RR | EDSS score of ≤ 4 | Not reported | N/A | 2x/week 30-40 minute sessions for 4 weeks | reflexologist (first author, who is the qualified reflexologist) | private room | waiting list control | No | MFIS | ISI | baseline, 2 months (appears there were other assessments time points but did not report data at those points) | There was no significant improvement in ISI between the intervention group (change mean 2.0, SD 4.7) and control group (change mean 0.8, SD 5.4; p = 0.146). |

Supplementary Table. Comprehensive data extraction. ACC = Active Control Condition; AE = Aquatic/Aerobic Exercise; BE = Basic Education; EDSS = Expanded Disease Status Scale; FOSQ-10 = Functional Outcomes of Sleep Questionnaire; FTC = Fatigue: Take Control; HEP = Home Exercise Program; HYP = Hypnosis; IAYT = International Association of Yoga Therapists; ICRP= Integrated Cognitive Rehabilitation Program; MAE = Moderate-Intensity Aerobic Exercise Program; MBCT = Mindfulness Based Cognitive Therapy; MBSR = Mindfulness Based Stress Reduction; MCID = Minimal Clinically Important Difference; MFIS = Modified Fatigue Impact Scale; MM = Mindfulness Meditation; MSFC = Multiple Sclerosis Functional Composite; MSTC = Multiple Sclerosis: Take Control; NHS = National Health Service; PDDS = Patient Determined Disease Steps; PHQ-9 = Patient Health Questionnaire; PMRT = Progressive Muscle Relaxation Technique; PP = Primary Progressive [Multiple Sclerosis]; PT = Physical Therapist; REM = Rapid Eye Movement; RN = Rest Night; RR = Relapse-Remitting [Multiple Sclerosis]; SE = Sleep Efficiency; SEFT = Supportive Emotion-Focused Therapy; SLP = Speech-Language Pathologist; SP = Secondary Progressive [Multiple Sclerosis]; SSES = Sleep Self-efficacy ScaleSSE = Sleep Self Efficacy; TAU = Treatment as Usual; TIB = Time in Bed; TN = Training Night; TRE = Trauma Releasing Exercises; TST = Total Sleep Time; WS = Walking and Stretching program.
